# Increasing dengue outbreaks in temperate Brazil is linked to *Aedes aegypti* invasion and infestation level driving widespread virus transmission

**DOI:** 10.1101/2025.02.24.25322773

**Authors:** Thales Bermann, Tatiana Schaffer Gregianini, Fernanda Marques Godinho, Ludmila Fiorenzano Baethgen, Fernanda Letícia Martiny, Erica Bortoli Möllmann, Amanda Pellenz Ruivo, Franciellen Machado dos Santos, Caroline do Nascimento Ferreira, Gustavo Barbosa de Lima, Danielly Alves Mendes Barbosa, Filipe Zimmer Dezordi, Milena Bauermann, Taina Machado Selayaran, Elgion Lucio da Silva Loreto, Felipe Gomes Naveca, André Luiz Sá de Oliveira, Ana Beatriz Gorini da Veiga, Marcelo Henrique Santos Paiva, de Lima Tulio Campos, Gonzalo Bello, Matheus Filgueira Bezerra, Felipe Valter de Oliveira, Rafael Maciel-de-Freitas, Tiago Gräf, Richard Steiner Salvato, Gabriel Luz Wallau

## Abstract

Dengue fever, caused by the dengue virus (DENV), is the most important mosquito-borne disease impacting human health globally, it is particularly prevalent in tropical regions where *Aedes* vectors thrive. However, due to a recent expansion of the main dengue vectors to subtropical and temperate zones in the last decades there is an increasing human population at risk. Brazil is a global hotspot for dengue, accounting for 70% of all dengue cases reported worldwide in 2024. Noteworthy, dengue became endemic even in the southernmost Brazilian state of Rio Grande do Sul, which until recently had only imported cases. In this study, we integrated entomological, epidemiological, and high-resolution genomic data to investigate DENV transmission dynamics in Southern Brazil over nine years. From 2015 to 2023, we detected a partial to full invasion of the state municipalities by *Aedes aegypti*, with dengue cases increasing substantially from 2020 onwards. The suitability index for transmission effectively predict the intensity of the outbreaks in the region, providing a two-month lead time before the surge in cases, representing an interesting proxy to forecast the outbreak size in the next few years. Invaded municipalities that surpassed a given case incidence level also increased, and by 2023, nearly half of the municipalities of the state surpassed that threshold, suggesting that dengue became endemic in the region. By analyzing 665 newly generated DENV genomes, we observed persistent overwintering lineages driving seasonal outbreaks, with the Northwest and Metropolitan regions serving as the main transmission hubs for dengue spread in the state. These regions were also the first to experience the endemic establishment of dengue lineages, making them key areas for targeted control efforts to mitigate further local outbreaks and the virus spread to other regions. These findings underscore the expansion of DENV-affected areas and increasing outbreaks into temperate zones, likely driven by the spread of the vector, associated with increased infestation level of the vector and widespread viral transmission in an immunologically naive population. In this context, enhanced control efforts are critical to mitigate the dengue burden in previously non-endemic areas.

## Introduction

Dengue fever, caused by the dengue virus (DENV), is one of the most significant mosquito-borne viral diseases affecting humans worldwide. In recent decades, dengue has evolved from a sporadic illness to a significant global public health concern, with a rapidly increasing incidence in tropical and temperate regions ^1^. The virus is primarily transmitted by *Aedes* mosquitoes, particularly *Aedes aegypti* in urban areas and *Ae. albopictus* in peri-urban settings ^2,3^. These mosquitoes predominantly grow in tropical regions worldwide, where they breed in both artificial and natural water reservoirs ^4^. Dengue fever is caused by four viral serotypes (DENV-1 to DENV-4), each likely originating from independent spillover events ^5^. Each serotype comprises multiple genotypes, and a variety of distinct lineages has recently been proposed to improve classification (Hill et al. 2024). Immunity is serotype-specific, and secondary infections are likely to increase the risk of severe disease, including dengue hemorrhagic fever (DHF) and dengue shock syndrome (DSS) ^6^.

Expansion of the dengue vector to new, previously unaffected subtropical and temperate areas, is expected to increase as a result of climate change and environmental degradation. Global warming is altering the distribution of vector-borne diseases, with regions previously considered non-endemic and low-risk for dengue transmission becoming increasingly vulnerable ^7–9^. Niche modeling studies have predicted a significant expansion in the *Aedes* mosquitoes habitat due to temperature rising ^10,11^. In addition, deforestation, land use, urbanization and other factors contribute to the increase in dengue and other arboviral diseases ^12,13^. Although many modeling studies forecast changes in the geographical distribution of vector-borne diseases, there are still limited reports on the impact of this phenomenon in the current epidemiology of dengue across the globe. Therefore, understanding the dengue virus transmission dynamics in previously non-endemic regions can provide not only clear examples of expanding diseases due to climate change but also essential information to guide targeted mitigation efforts, ultimately reducing the burden of dengue in subtropical and temperate regions worldwide.

Brazil has emerged as a global dengue hotspot, consistently reporting the highest number of cases worldwide; with 10 million cases reported in 2024, Brazil accounted for 70% of the total global dengue cases in that year ^14^. The Brazilian vast and diverse geography, along with its tropical climate and unplanned urban development, makes the ideal environment for *Aedes* mosquitoes ^15^. Moreover, dengue is now becoming more common in previously non-endemic Brazilian areas, attributed to the increasing average temperature and, consequently, the spread of the virus to previously unexposed populations ^16^. Rio Grande do Sul, the Brazilian southernmost state, has historically been less affected by dengue compared to the tropical regions of the country ^17^. However, this scenario has dramatically changed in recent years ^18^, as the state has experienced recurrent outbreaks, reaching over 207,000 dengue cases in 2024 ^19–21^.

In this study, we used mosquito population, epidemiological data, and high-resolution genomic data to uncover the DENV transmission dynamics in Southern Brazil over nine years (2015–2023). Our findings revealed that *Ae. aegypti* has now spread completely throughout the state, driving increasing outbreaks of overwintering dengue lineages. Moreover, we uncovered the two main dengue transmission hubs in the state. Taking into consideration the expected increasing dengue burden in the region in the next few years, and that outbreak intensity can be forecasted by the DENV transmission suitability index associated with known dengue transmission hub hotspots, there is now the possibility of targeted efforts to mitigate human suffering and deaths in the region.

## Methods

### Historical dengue epidemiology

To assess the invasion and transmission dynamics of dengue fever cases in Rio Grande do Sul, Brazil, we conducted an invasion modeling analysis following the approach of Harish et al. (2024). Based on historical records of dengue incidence in Brazil, these authors defined an invaded municipality as one that surpassed the annual incidence of ≥20 cases per 100,000 residents per year. These thresholds were chosen to balance the identification of areas that, once invaded, consistently report cases and can act as dispersion hubs. For this analysis, we retrieved dengue case data from Rio Grande do Sul from 2015 to 2023 at the municipality level from the Notifiable Diseases Information System (SINAN) platform ^22^. Population size data for each municipality were obtained from the most recent national census conducted by the Brazilian Institute of Geography and Statistics (IBGE) in 2022 ^23^. Using this information, we calculated annual dengue incidence rates for each municipality.

Dengue incidence has been historically concentrated in the Northwest and Metropolitan regions of the state. To better understand the spatial distribution of dengue cases, we considered the division of Rio Grande do Sul into its seven subregions, as defined by the IBGE: Northwest, Northeast, Metropolitan, Center-East, Center-West, Southeast, and Southwest. These subregions provide a broader geographic framework for analyzing dengue invasion patterns and transmission dynamics across the state. The incidence rates were then visualized using thematic maps generated with the *Tmap* package in R ^24^, allowing us to spatially depict variations in dengue incidence across the state and identify invaded municipalities that meet the criteria for endemic transmission as defined by Harish et al. (2024).

### Historical mosquito population estimates

*Ae. aegypti* monitoring was implemented in Rio Grande do Sul in 2015 by the State Center for Health Surveillance (CEVS), and the data are publicly available at https://ti.saude.rs.gov.br/dengue/painel_de_casos.html ^25^. According to this surveillance dataset, a municipality is classified as infested when *Ae. aegypti* immature (larvae or pupae) are found in or near households. The infestation status was treated as a binary variable (0 = not infested; 1 = infested), and data were collected from 2015 to 2023, covering the study period. The spatial distribution of infestation was visualized using maps generated with the Tmap package in R. As a complementary analysis, we incorporated an additional dataset from the Brazilian Ministry of Health, which provides a more detailed classification of infestation levels in Rio Grande do Sul ^26,27^. This dataset categorizes infestation into four levels: 0 (absence of infestation), 1 (low infestation), 2 (moderate infestation), and 3 (high infestation); however, this dataset only covers the period from 2018 to 2023. The integration of both datasets enables a more comprehensive evaluation of *Ae. aegypti* infestation trends in the region.

### Climate and environmental suitability based on Index P

The mosquito-borne viral suitability index (*Index P*) was estimated using the MVSE package in R ^28^. This index quantifies the potential for pathogen transmission by mosquitoes, mathematically assessing how climatic factors such as temperature, humidity, and precipitation influence the generation of new infections. The estimation was performed on a daily scale for the state of Rio Grande do Sul, covering the period from January 2015 to December 2023. Climatic data, including daily mean temperature, daily mean humidity, and daily precipitation, were obtained from the Brazilian Meteorological Database for Teaching and Research (BDMEP) ^29^, considering both automatic and conventional weather stations that recorded these variables within the studied area. A total of 35 municipalities with available climate records were selected. For days with missing climate data, gaps were filled using averages calculated from corresponding days and months of other years. Once the dataset was completed, we applied the Inverse Distance Weighting (IDW) interpolation method, implemented through the scipy.spatial.distance module in Python ^30^, to estimate temperature and humidity values for municipalities without direct meteorological station coverage.

### Statistical analysis

Given the non-parametric distribution of yearly incidence rates per municipality, we used the Mann-Whitney test to assess whether the mosquito infestation status was associated with increased dengue incidence within the same year at the municipality level. We used exponential correlation tests to explore the correlation between P index and dengue incidence at both the subregion and state levels. Receiver Operating Characteristic (ROC) curves were used to evaluate the performance rates for the P index as predictor for epidemics months. Based on the exponential correlation between incidence and P index, we calculate the slope of each year equation (K values).

Then we used linear regression to test the hypothesis of modulation of the K values by the dissemination of DENV in the territory, using the fraction of total territory area with confirmed cases (sum of the areas from positive municipalities by subregion) as a proxy for DENV dissemination. All statistical tests were performed with a 95% confidence interval. Statistical tests and graphs were performed using GraphPad Prism version 8.

### Patient samples and DENV detection

This study included 8,951 serum samples from individuals with suspected acute febrile illness, collected across 130 cities in the state (out of 497 cities total) between 2015 and 2023 and received by the State Public Health Laboratory of Rio Grande do Sul (LACEN-RS) for DENV detection and serotyping. Ribonucleic acid (RNA) was isolated from the samples using automated extraction on the Extracta 96 equipment (Loccus), utilizing the Kit Fast Viral DNA and RNA (MVXA-P096 FAST) (Loccus, São Paulo, Brazil), according to the manufacturer’s instructions. DENV detection and serotyping were carried out through quantitative reverse transcription polymerase chain reaction (RT-qPCR) using the Molecular ZDC Kit (Bio-Manguinhos, Brazil), following the manufacturer’s instructions, and performed on ABI 7500 Real-Time PCR System (Applied Biosystems, Foster City, CA, USA) and CFX Opus Real-Time PCR System (Bio-Rad, Hercules, CA, USA). Samples that showed amplification with a cycle threshold (Ct) ≤38 were considered positive for DENV. Positive and negative controls were included in each analysis.

### Genome sequencing and assembly

A total of 3,984 DENV-positive samples were identified by RT-qPCR at LACEN-RS over the studied period. We obtained raw reads from 626 samples from DENV-1 infection cases and 39 from DENV-2, totaling 665 samples investigated through whole genome deep sequencing. Genomic libraries were prepared using an adaptation of the Illumina COVIDseq protocol (Illumina, San Diego, USA), replacing the SARS-CoV-2 primers with previously published DENV primer schemes ^31,32^. Sequencing was performed using the Illumina Miseq platform (Illumina, San Diego, USA.). A reference-based genome assembly was conducted using the ViralFlow 1.0.0 pipeline ^33^, using the closest and well-annotated DENV-1 (GenBank: GU131863.1) and DENV-2 (GenBank: MW577818.1) reference genomes. The average coverage breadth across the genome from all samples ranged from 72.2% to 99.8%, and an average coverage depth of 1817.46 (SD=908.59), ranging from 294.0 to 6,625.5 (Supplementary Table 1).

### Genotyping and phylogenetic analyses

To assign DENV genotypes and lineages to each sequence, we used the recently introduced dengue lineage system defined by Hill et al. (2024) (dengue-lineages.org), and implemented in the Genome Detective Virus Tool Version 2.72 ^34^. To characterize the transmission chains of DENV lineages, we downloaded all near-complete genomic sequences of DENV-1 and DENV-2 with coverage >70% available at the NCBI virus database (accessed in March 2023). Our dataset included 3,476 DENV-1 and 3,091 DENV-2 publicly available sequences from NCBI. The retrieved sequences were then aligned with those generated in this study using MAFFT ^35^, and manually curated to remove artifacts using AliView ^36^. Maximum likelihood (ML) phylogenetic trees were estimated using IQ-TREE2 ^37^ under the General Time Reversible (GTR) nucleotide substitution model, which was inferred as the best-fit model by the ModelFinder application implemented in IQ-TREE2 ^38^. Statistical support for tree nodes and branches was estimated using ultrafast bootstrap approximation (UFboot) and approximate likelihood-ratio test (aLRT) approaches under 1000 replicates ^39,40^. From this complete dataset, we extracted subsets with the identified lineages and clades for further subsequent analyses.

### Phylodynamics analyses

Our phylodynamic analyses were conducted separately for eight subsets, which were defined based on the ML phylogenies of global DENV-1 and DENV-2 datasets. Each subset composition is detailed in Supplementary Table 2 and Figure S1. These subsets were then used individually for time-calibrated phylogenetic analyses and spatial diffusion reconstruction. To assess the temporal signal in our subsets, we performed a root-to-tip regression analysis between divergence from the root and sample collection dates in each ML tree using TempEst v.1.5.1 ^41^. The results were exported to a table and subsequently analyzed via an in-house R script to identify outliers using the interquartile range by the Tukey method ^42^. To estimate time-calibrated phylogenies, we conducted a phylogenetic analysis for each subset individually using a Bayesian approach with BEAST v1.10.4 ^43^. We employed the SDR06 codon-based substitution model ^44^, which is well-suited for viral datasets as it explicitly accounts for differences in evolutionary constraints among codon positions, providing greater flexibility in modeling the increased substitution rate at third codon positions. Path Sampling (PS) and Stepping-Stone Sampling (SS) scores ^45,46^ of different molecular clock models with different demographic models were assessed for each subset to determine the most suitable demographic and molecular clock models.

Initially, we performed demographic model selection by running a Bayesian skyline plot as the demographic model. This approach allowed us to estimate the time to the most recent common ancestor (tMRCA) for each subset and ensured sufficiently high Effective Sample Size (ESS) values. Based on the results, we used the tMRCA estimates obtained from the skyline plot as input for setting the "time at last transition point" when testing the SkyGrid model. The SkyGrid model achieved the best fit among the demographic models, as evidenced by its superior PS and SS scores in a marginal likelihood estimation (MLE) analysis (Baele et al. 2012, 2013). We proceeded with the SkyGrid model for downstream analyses. Supplementary Table 3 provides details on the ’time at last transition points’ and the number of grids defined per year for each subset, while Supplementary Table 4 presents the path sampling and stepping-stone values for each combination of clock model and demographic model tested. Analyses were conducted under 200 million MCMC steps, sampling parameters and trees every 20,000 steps. Convergence of MCMC chains was checked using Tracer ^47^. Maximum clade credibility (MCC) trees for each subset were summarized using TreeAnnotator after discarding 10% as burn-in. Spatial diffusion reconstruction was performed on the posterior sampling of the trees using a discrete model with asymmetric substitution rates and Bayesian stochastic search variable selection inference (BSSVS), conducted over 10 million MCMC steps ^48,49^. To visualize the results, we used the SPREAD 4 tool ^50^; for each discrete location, coordinates of latitude and longitude were attributed.

## Results

### Historical dengue epidemiology in Rio Grande do Sul

The South Region of Brazil comprises the states of Paraná, Santa Catarina and Rio Grande do Sul Figure 1A. The distribution of dengue cases in this region has been influenced by various environmental factors, including the distinct climatic zones present in the area. As shown in Figure 1A, there is a transition from the tropical climate zone that predominates in Brazil, which extends into the northern part of Paraná, to the temperate climate zone that characterizes the southern portions of the South Region. Between 2015 and 2023, Rio Grande do Sul recorded 214,695 suspected cases of dengue, of which 125,500 were later confirmed by molecular assay. The annual distribution of confirmed dengue cases, illustrated in Figure 1B, shows the highest number of cases in 2022, with 67,345 reported cases, and the lowest in 2017, with 26 cases ^21^.

**Figure 1.**
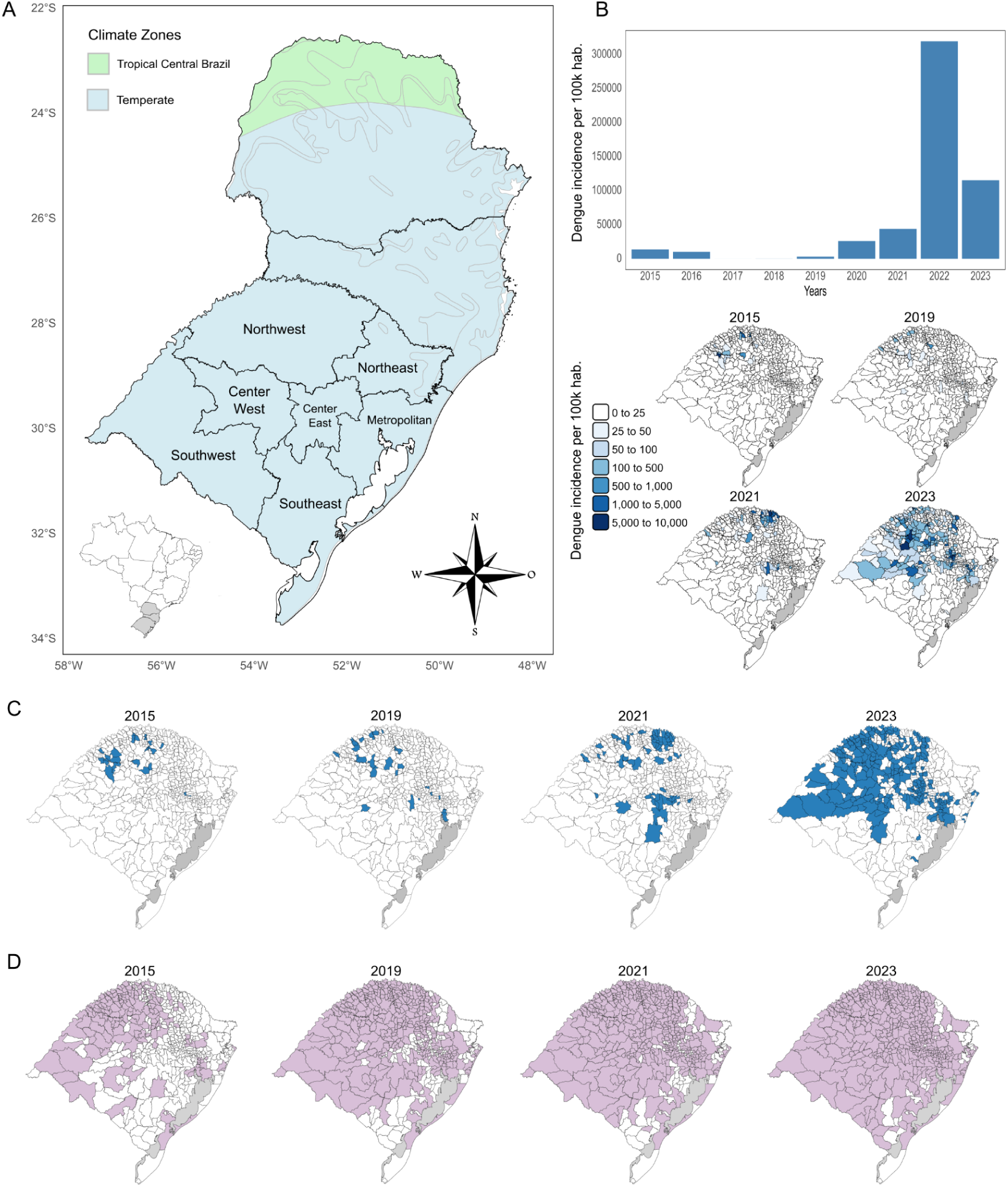
Historical spatiotemporal dengue epidemiology in Rio Grande do Sul. (A) Reference map of Brazil with a zoomed-in view of the southern region, covering the three states: Paraná, Santa Catarina, and Rio Grande do Sul. Rio Grande do Sul, the southernmost state, is depicted with the subregions of the state (IBGE, 2020). Climate zones are depicted in green (tropical) and blue (temperate). (B) The upper panel depicts the incidence of dengue cases (per 100,000 inhabitants) in Rio Grande do Sul during the study period (2015–2023); the lower panel shows the incidence per municipality in the years 2015, 2019, 2021, and 2023. (C) Municipalities invaded by DENV (incidence ≥ 20 cases/100,000 inhabitants) during the study period. (D) Municipalities invaded by *Aedes aegypti*.

Dengue incidence in Rio Grande do Sul increased 17.36-fold when comparing 2015–2019 to 2020–2023 (Figure 1B upper panel). In recent years, the incidence has increased in several regions of the state including the ones with comparatively lower incidence such as: Center-West, Center-East, and Southwest sections as well (Figure 1B lower panel). We also mapped the dengue-invaded municipalities (>20 cases/100,000 inhabitants) and found an increase from 22 municipalities in 2015 to 254 in 2023 (Figure 1C), with a higher predominance of invaded municipalities in the Northwest, Center-West, Center-East, Southwest, and Metropolitan portions of the state (Figure 1C).

### Historical mosquito population invasion

Based on the surveillance data available on the CEVS platform, our analysis found that *Ae. aegypti* had already infested nearly half of the municipalities in Rio Grande do Sul by 2015, with the regions with highest infestation levels being the Northwest, Center-West, and Southwest regions. In contrast, the Metropolitan, Northeast, Center-East, and Southeast regions had only a few invaded municipalities in 2015 (Figure 1D). In the following years, infestation spread rapidly, reaching almost all municipalities of the state by 2023 (Figure 1D).

To further characterize the progression of *Ae. aegypti* infestation levels in Rio Grande do Sul, we analyzed data from the Brazilian Ministry of Health. The results indicate a gradual increase in infestation levels across municipalities over the years. In 2018, about 36% of municipalities were classified as having no infestation (category 0), 25% had low infestation (category 1), and only 4% had moderate infestation (category 2). Starting in 2019, there was a progressive reduction in municipalities with no infestation, dropping to 33% in 2023, while higher categories increased. The highest infestation level (category 3) was first detected in 2022, and in 2023, its occurrence persisted. That year, 24.7% of municipalities had low infestation, 21.5% had moderate infestation, and 7.9% had high infestation – this being the first time the highest category was recorded in consecutive years.

The *Aedes aegypti* infestation index (LIRA – Levantamento Rápido de Índices do *Aedes aegypti*) and dengue incidence was analyzed as shown in Supplementary Figure X, municipalities with infestation levels 1, 2, and 3 had significantly higher average dengue incidence (2018–2023) compared to those with no detected infestation (p < 0.0001). Dengue incidence was also significantly higher in municipalities with high infestation (Lira 3) compared to those with moderate infestation (Lira 2) (p = 0.0014) Figure S4.

### Climate and environmental suitability for dengue virus transmission

We used the mosquito-borne viral suitability index to assess the temporal and spatial variation across different regions within Rio Grande do Sul state (Figure 2). The results revealed a clear seasonal pattern, with high suitability periods typically occurring during the warmer months of each year (January to March). No increase was detected in either the peak values of Index P or the length of periods with consecutive months where Index P exceeded 1 during the analyzed years. The complete results of Index P for each state region are presented in Figures S5 and S6.

**Figure 2.**
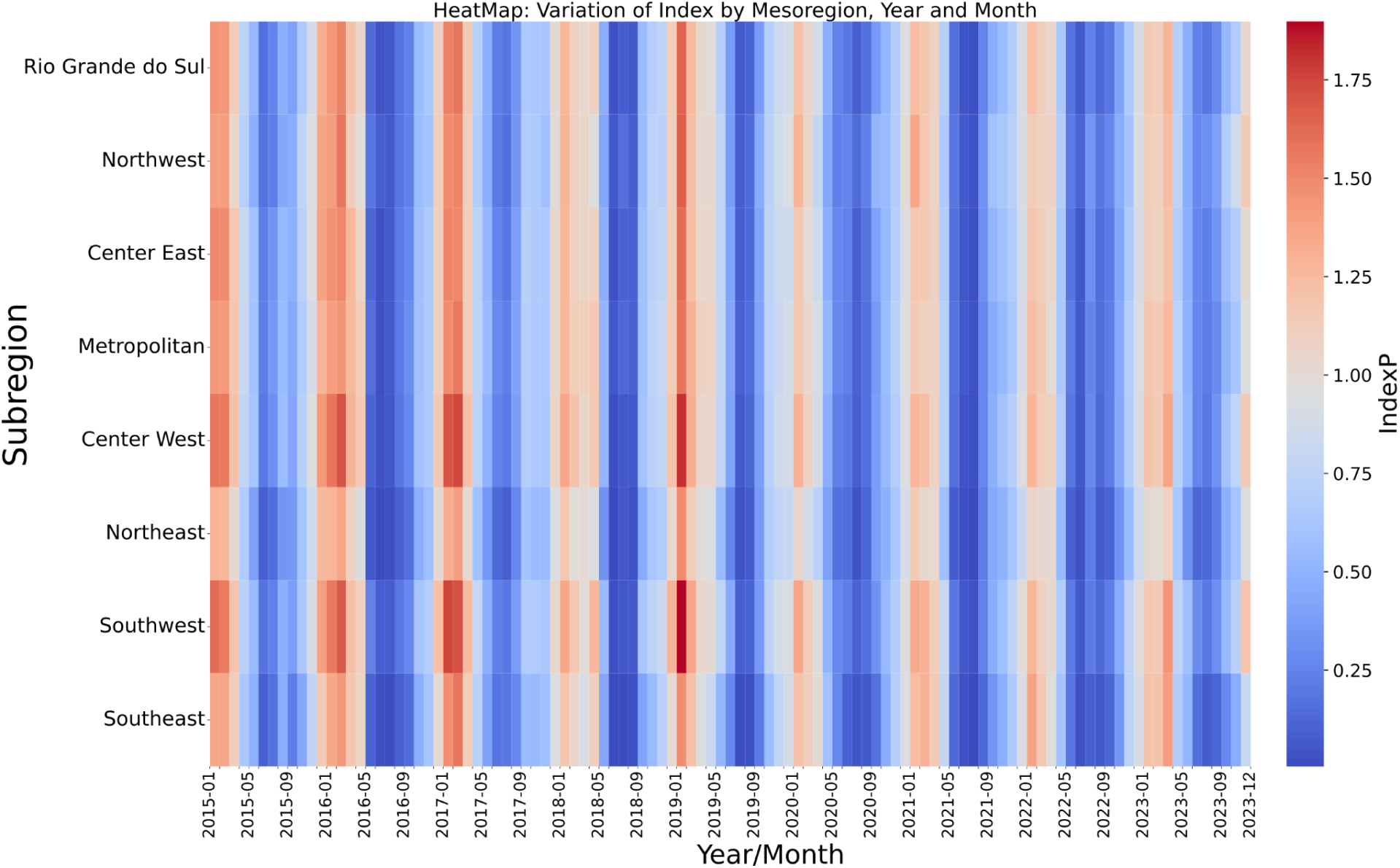
Mosquito-borne viral suitability in Rio Grande do Sul state. The mosquito-borne viral suitability index (Index P) was estimated for the Rio Grande do Sul state from January 2015 to December 2023. The color scale represents the intensity of the index, with red shades indicating higher transmission potential (Index P >1) and blue shades representing lower values. The figure highlights the temporal evolution of transmission risk across different regions, showing an increasing persistence of high-risk periods over time, particularly in the Northwest, Center-West, Southwest, and Metropolitan regions.

Of note, the mosquito-borne viral suitability index (P index) exhibited a strong positive exponential correlation with the number of dengue incidence per year (Figure 3A). However, this correlation was only observed when a two-month lag was considered, with the strength of the association varying across years (Figure 3B). Consistently, ROC curve analysis demonstrated that the two-month lagged P index, but not the concurrent-month index, was predictive of epidemic months (sensitivity = 93.5, CI95% = 79.2-98.8; specificity = 84.0, CI95% = 74.1-90.6; Figure 3C, Supplementary table 5). This exponential relationship was evident at both state and subregional levels, with dengue incidence showing temporal and spatial heterogeneity. In 2022, only four subregions exhibited a high correlation, while three others (Center West, Southwest, and Southeast) maintained a moderate association. By 2023, however, this scenario shifted, with a strengthening of the correlation across nearly all subregions, indicating a more consistent relationship between the variables analyzed.

**Figure 3.**
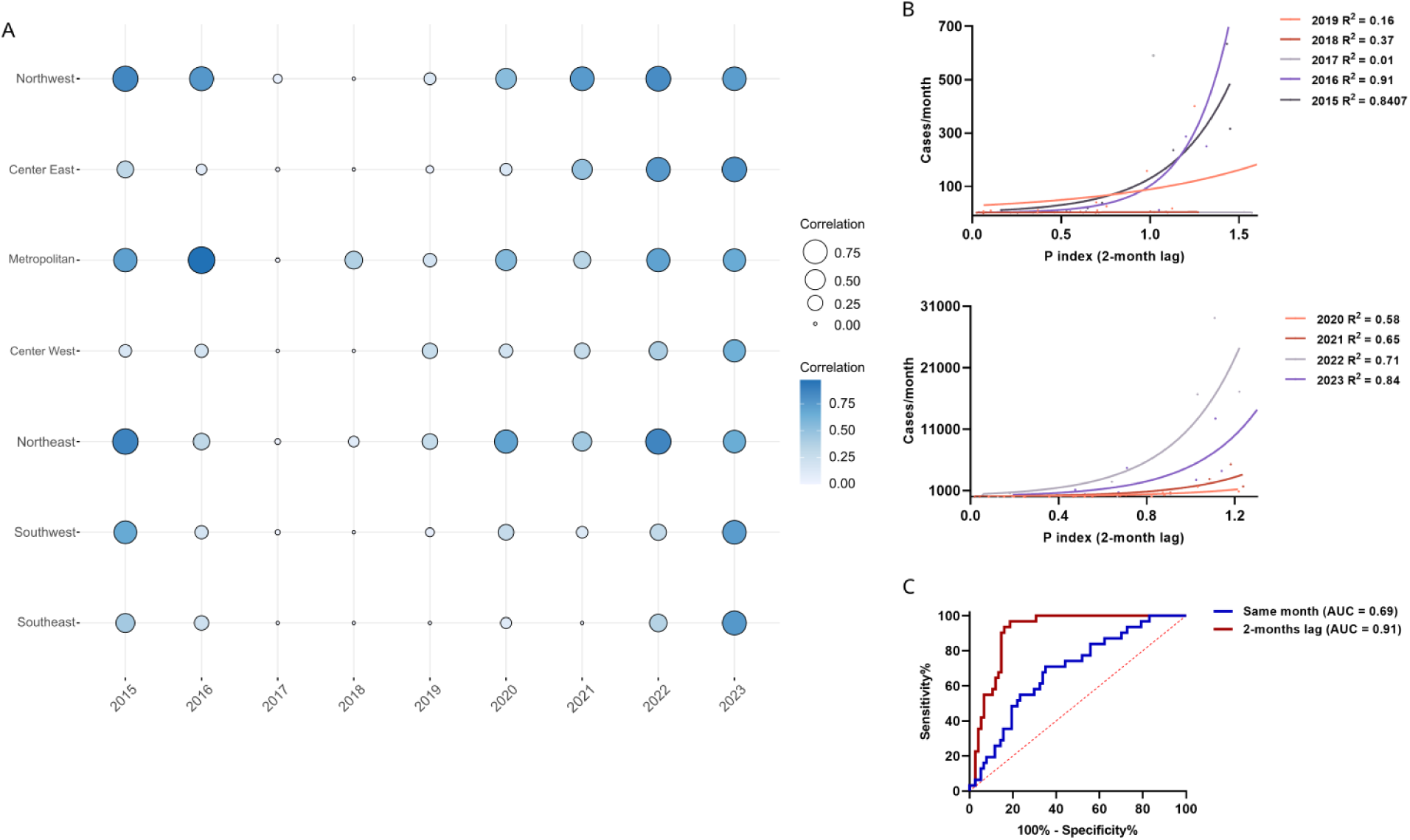
Correlation between the mosquito-borne viral suitability index (P index) and dengue cases in Rio Grande do Sul. (A) Correlation (R²) between dengue incidence and P index according to the mesoregions, between ion from 2015 to 2023, with bubble size and color indicating correlation intensity. (B) Positive exponential correlation between monthly dengue cases (Y-axis) and the P index with a two-month lag (X-axis), aggregated by year - 2015-2019 and 2020-2023 periods. (C) ROC curve demonstrating the predictive performance of the lagged and unlagged P index for epidemic months.

Interestingly, the slope of the confirmed cases/P index correlation increased over the study period, as represented by the K constant, suggesting the influence of an additional factor on dengue cases (Figure 4A). Given that the study period coincided with the progressive introduction of DENV into a previously Dengue-naïve region, we hypothesized that the extent of viral geographical spread might also contribute to the incidence dynamics, co-dependently with the P index. To test this hypothesis, we used the proportion of the total area of dengue-positive municipalities within a subregion in a given year as a proxy for viral spread, referred to as the "area fraction."

**Figure 4.**
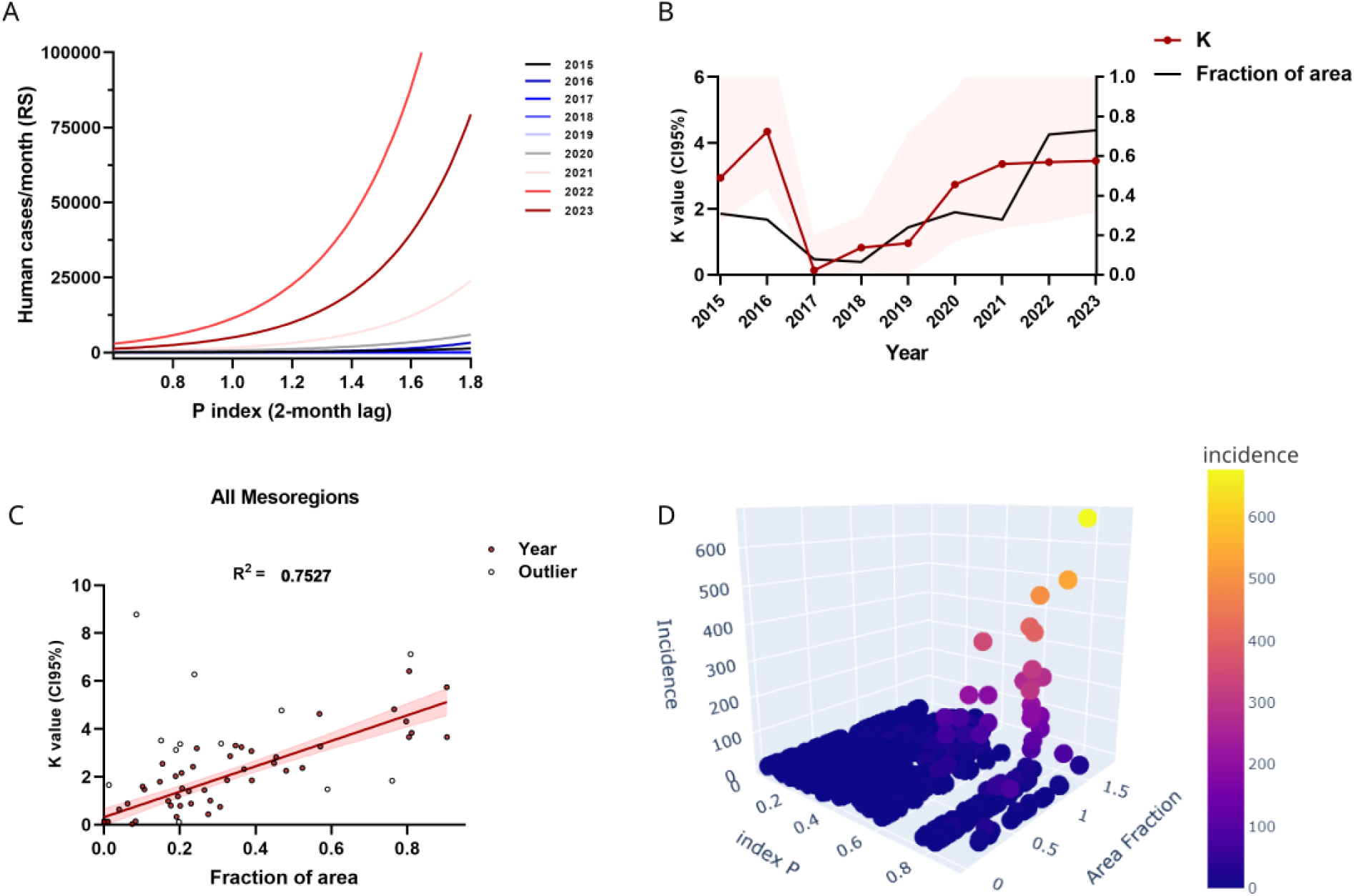
Role of the fraction of area with confirmed cases on the P index/incidence correlation. (A) Shift in the slope of the exponential curve from the P index/confirmed cases in RS. (B) on the left Y axis, the increase of the K constant, which determines the exponential slope from Figure A. On the right Y axis, a similar increase of the fraction of the area represented by dengue-positive municipalities was observed. (C) Linear regression of the K values in function of the área fraction. Each data point represents the values from one mesoregion in a given year. (D) three dimensional scatterplot showing the co-dependency of dengue incidence with the lagged P index and area fraction with positive municipalities.

Linear regression analysis revealed that the K constant (slope of the incidence/P index correlation) exhibited a direct linear relationship with the annual area fraction of dengue cases (Figures 4B, C and D). Together, these findings suggest that the P index and the yearly dengue area fraction have the potential to be further explored as predictive tools for estimating case incidence two months in advance.

### Genomic diversity of dengue virus in Southern Brazil

In this study, genomic sequences of DENV were successfully obtained from 665 samples, including 626 of DENV-1 and 39 of DENV-2 serotypes, which were state-wide representative of most regions (Figure 5A). Genotyping analysis assigned all 626 DENV-1 sequences into genotype V, lineages 1V_E, 1V_D.1, 1V_F, and 1V_A (Figure 5B). Most of the DENV-1 sequences were assigned to the 1V_E lineage (54.0%), followed by 1V_D.1 (35.3%), 1V_A (6.4%), and 1V_F (4.3%). The DENV-1V_A lineage was dominant from 2015 to 2016; by 2021 to 2023, the 1V_D and 1V_E lineages had become the most prevalent. The 1V_F lineage initially detected in 2015, reappeared in 2022. Among the 39 DENV-2 sequences, 32 (82.1%) were identified as genotype II – Cosmopolitan (lineage 2II_F.1.2), while seven (17.9%) were classified as genotype III – Southeast Asian/American (lineage 2III_C.1.1) (Figure 5B).

**Figure 5.**
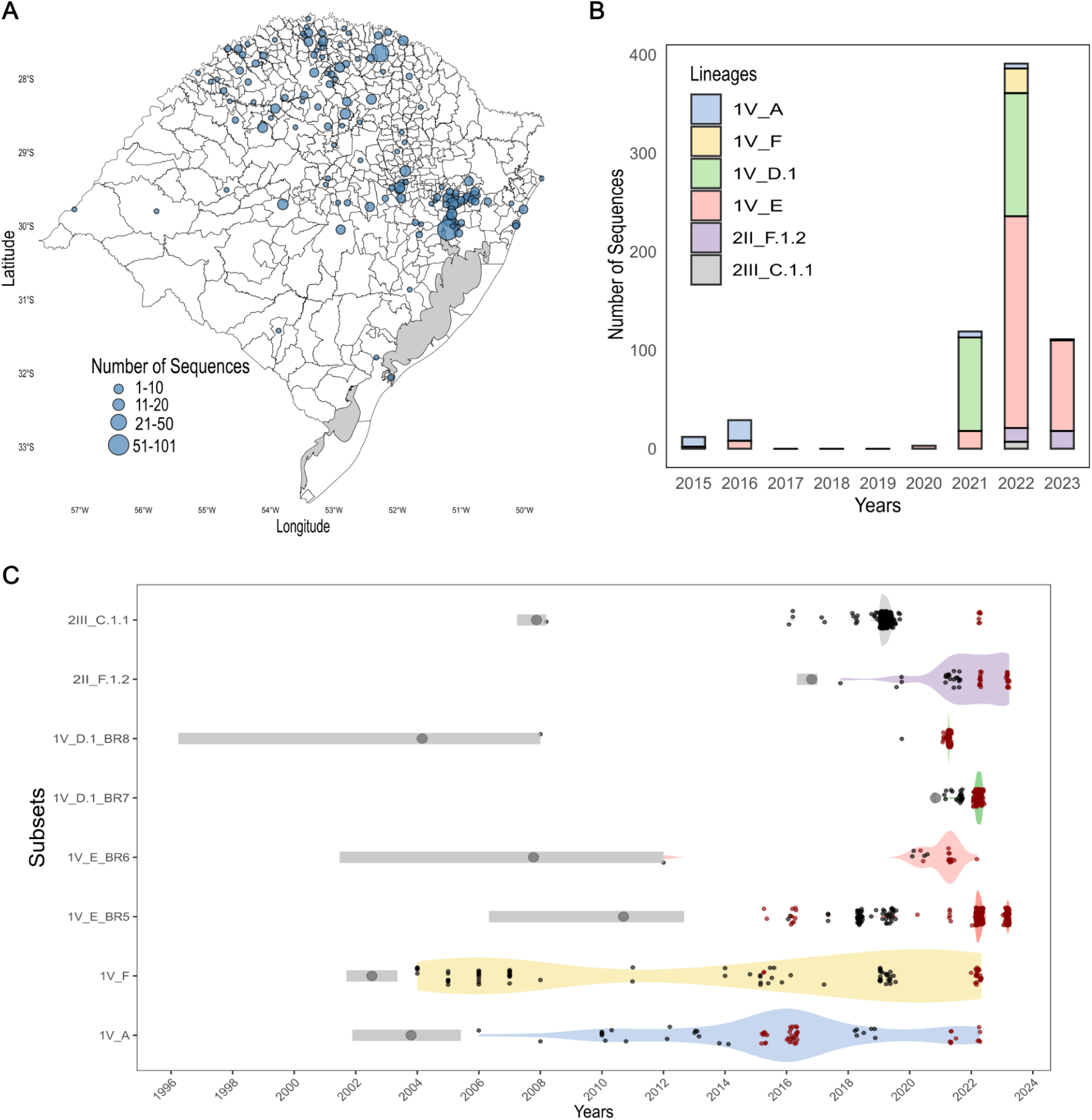
Spatiotemporal distribution of DENV lineages in Rio Grande do Sul (2015-2023). (A) Rio Grande do Sul state map showing the geographic locations of the 665 DENV samples sequenced in this study (626 DENV-1 and 39 DENV-2). (B) Time series of DENV sequences generated by year according to the assigned lineages. (C) Height posterior density of the tMRCA for each clade. The violin plots represent the temporal distribution of sampled sequences, with wider sections indicating a higher concentration of sampled genomes over time. Red dots represent sequences generated in this study, while black dots indicate publicly available sequences from the NCBI database. In parallel, the gray bars denote the estimated time to the most recent common ancestor (tMRCA) for each subset, with the length of the bar representing the 95% highest posterior density (HPD) interval. The central circle within each gray bar marks the median tMRCA estimate. The x-axis spans from 1996 to 2024, providing a temporal reference for both sampling efforts and inferred lineage ancestry.

Phylogenetic reconstruction revealed the presence of two well-supported clades, previously named BR5 and BR6 ^31^ within the DENV-1V_E lineage. Similarly, the DENV-1V_D.1 lineage was subdivided into two well-supported clades that we named BR7 and BR8 (Supplementary Figure 1). Based on these identified lineages and clades, we extracted six subsets from the DENV-1 dataset and two subsets from the DENV-2 dataset, covering all Rio Grande do Sul DENV diversity in the studied period for subsequent phylodynamic analysis. The distribution of our sequenced samples among the different DENV lineages and clades, including a more detailed description of the assembled subsets, is presented in Supplementary Table 2.

### Spatiotemporal spread of dengue virus in Rio Grande do Sul

To investigate the spatiotemporal transmission dynamics of DENV in Rio Grande do Sul, we first confirmed the molecular clock signal across all subsets. Afterward, we implemented a relaxed uncorrelated clock alongside the Bayesian Skygrid demographic models to explore the phylodynamics of DENV in the region. For DENV-1 subsets, the tMRCA estimate ranged from July 2002 to November 2020, while for DENV-2 subsets, the estimated time ranged from November 2007 to October 2016 (Figure 5C). The DENV-1V_A lineage was the predominant lineage during the 2016 outbreak, while 1V_E was also present in 2016 but at a lower frequency. Lineages 1V_F, 1V_E, 1V_D.1 and 2II_F.1.2 were primarily associated with the 2022 outbreak in Rio Grande do Sul.

The DENV-1V_E lineage exhibited a change in spatial diffusion pattern over the studied period. Between 2012 and 2018, this lineage was introduced multiple times into the state, contributing to the 2016 dengue outbreak (Figure 6A). However, from 2019 to 2023, this lineage transitioned to a more endemic transmission, spreading from the metropolitan region to central and northwestern areas. This lineage was present in both the 2016 (small-scale) and 2022 (large-scale) outbreaks, though with distinct dynamics between the two events. As the predominant lineage in the dataset, 1V_E accounted for approximately 50% of the sequenced samples. The spatial diffusion dynamics of this lineage showed a progressive expansion from the metropolitan region, with multiple dispersal events toward the northwest between 2016 and 2023. From 2019 onward, additional introductions occurred into the central-east, followed by a more widespread movement toward the central-west and southern regions (southeast and southwest) between 2021 and 2023. These diffusion patterns highlight the shifting role of 1V_E over time, from repeated introductions to sustained local transmission across broader geographic areas in the state (Figure 6B).

**Figure 6.**
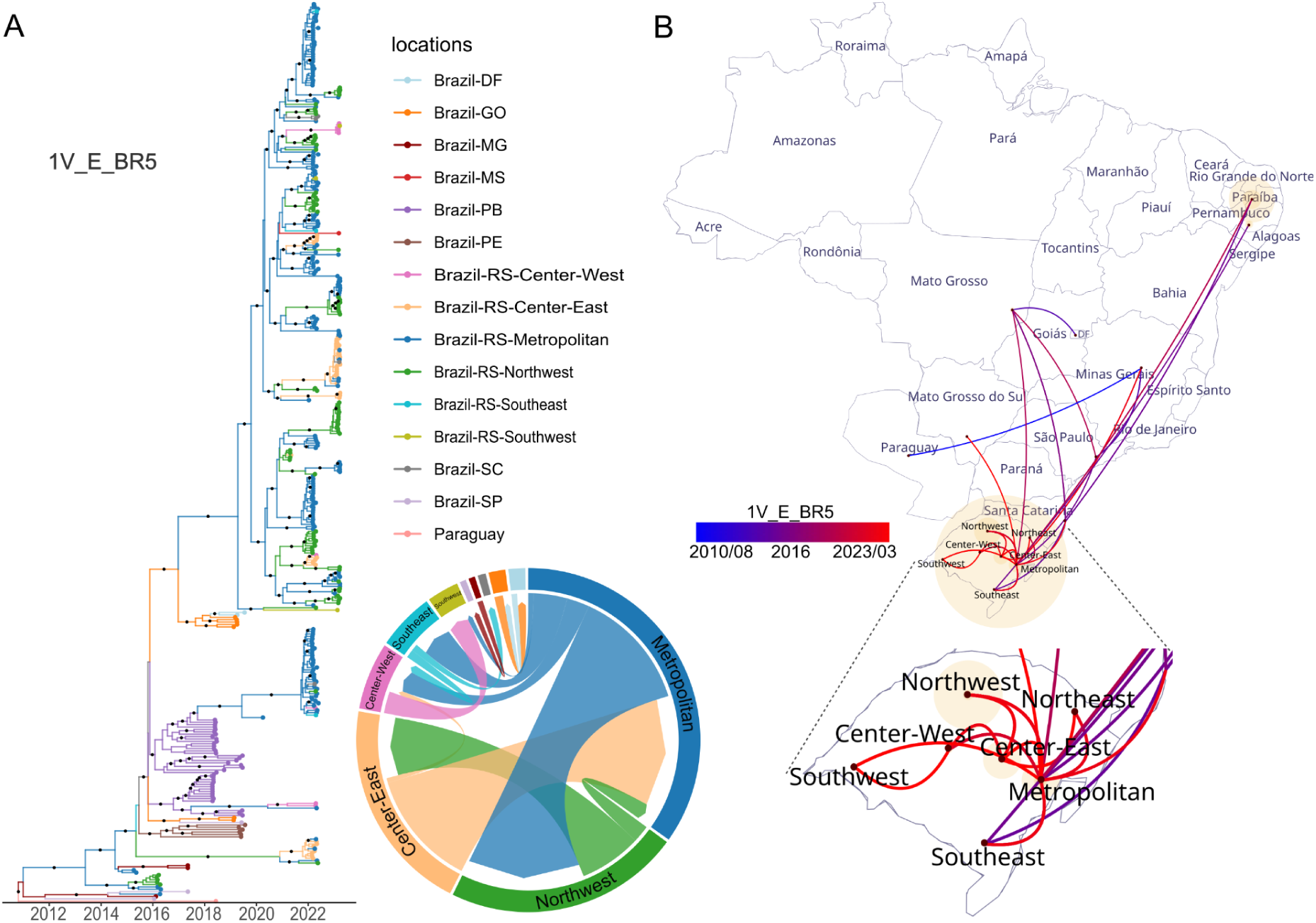
Spatiotemporal spread of the DENV-1V_E_BR5 lineage in Rio Grande do Sul. (A) Maximum clade credibility tree on a temporal scale for lineage 1V_E_BR5, illustrating its temporal evolution and geographic diffusion across localities. The colors correspond to the defined localities, and the circular migration plot represents the migration dynamics between these regions, as inferred from Markov jump analyses. (B) Spatial diffusion analysis depicting the inferred spread of lineage 1V_E_BR5 across different regions of the state. The lines represent dispersal events, with colors corresponding to different time periods. The inset highlights the diffusion patterns within Rio Grande do Sul.

In contrast, the DENV-1V_D.1 lineage represented an emergent lineage with a tMRCA estimated in November 2020. The first 1V_D.1 sequences were detected in Peru in early 2021, and the lineage was later identified in Rio Grande do Sul in late 2021, with two distinct dispersal patterns observed. Clade 1V_D.1_BR7 was introduced into the northwest region before spreading toward the metropolitan area (Figure S11), while clade 1V_D.1_BR8 arrived independently in the northwest but later dispersed into both the central-east and central-west regions between 2021 and 2022 (Figure S12). This lineage played a significant role in the 2022 outbreak, peaking during the dengue season (December to March) and comprising over 30% of the sequenced samples (Figure 7A).

**Figure 7.**
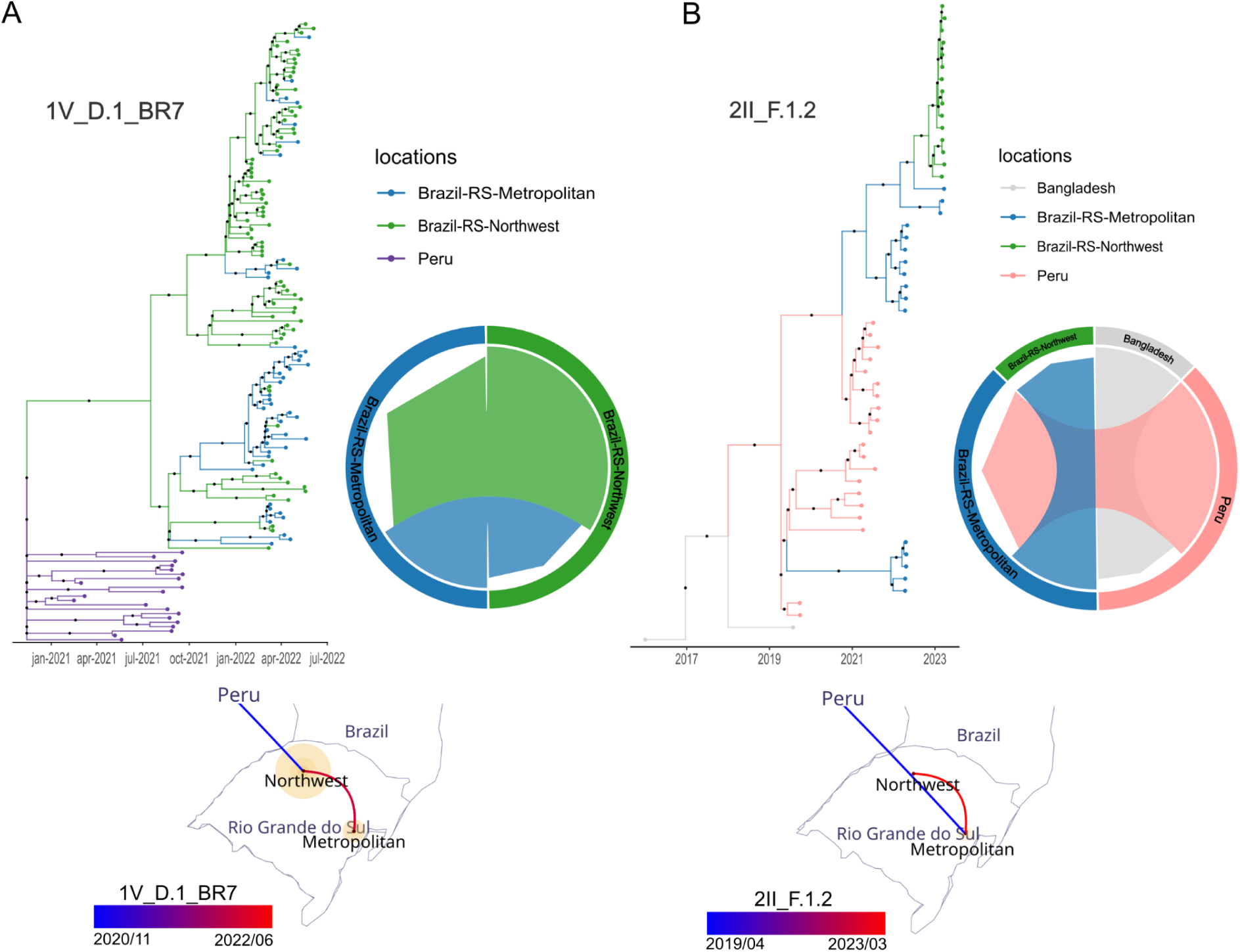
Spatiotemporal evolution of emergent DENV-1 and DENV-2 lineages. (A) Maximum clade credibility tree on a temporal scale for DENV-1V_D.1 lineage, with branch colors indicating different geographic regions. The circular migration plot represents the inferred migration dynamics between regions, while the spatial diffusion map illustrates the inferred spread of the lineage across Rio Grande do Sul. (B) Maximum clade credibility tree for DENV-2 lineage 2II_F.1.2, along with the corresponding circular migration plot and spatial diffusion map. The dispersal routes highlight the inferred geographic movements of the lineage between Peru and Rio Grande do Sul, as well as within Rio Grande do Sul.

Beyond these lineages, the DENV-1V_A lineage was introduced into the metropolitan region in 2011, subsequently dispersing to the northwest in 2015 and the central-west in 2016. While it played a role in the 2016 outbreak, its spread remained confined to that event, with only sporadic detections in 2021 and 2022 at low frequencies. Similarly, the 1V_F lineage first arrived in the metropolitan region in 2011, circulating at low levels before being detected in the dataset in 2015. A second introduction event in 2020 led to its establishment in the central-east, from where it expanded into the central-west in 2022 (Figure S7 and S8).

Among the DENV-2 lineages, 2II_F.1.2 accounted for 4.81% of the sequenced samples and represented a new genotype (Genotype II – Cosmopolitan) circulating in Brazil. Initially identified in Peru in 2019 and 2021, this lineage was subsequently detected in Rio Grande do Sul in 2022 and 2023 ^32^. Its emergence coincided with the large outbreak of 2022 and followed a dispersion pattern similar to that of 1V_E, 1V_A, and 1V_F, spreading from the Metropolitan region toward the Northwestern areas of Rio Grande do Sul (Figure 7B). Meanwhile, the DENV-2III_C.1.1 lineage was represented by only seven samples from 2022, indicating limited detection in the dataset. This lineage was first observed in the northwest and later dispersed into the central-west and central-east regions (Figure S14). Phylogeographic dynamics data for all eight subsets, including the MCC trees and circular migration plots from Markov jump analyses, are available in the supplementary material (Figures S7–S14). Additionally, interactive spatial diffusion maps for all lineages, generated using SpreaD4, can be accessed through the links provided in Supplementary Table 6. The well-supported regional transition events inferred from Markov jump analyses for each subset are detailed in Supplementary Table 7.

## Discussion

Ongoing climatic changes have substantially impacted rainfall patterns and temperature fluctuation across the globe, and significant displacement of species, vectors, and diseases is expected to increase in the coming years/decades ^51^. Vector-borne diseases are expected to be influenced by climate change due to the association between temperatures and humidity, which facilitates the establishment of mosquito vectors in new areas, thereby raising the burden of previously tropical arthropod-borne pathogens ^52^. Dengue is one of the best examples of an arbovirus that is expanding its range to subtropical and temperate areas due to the spread of the anthropophilic *Ae. aegypti* and *Ae. albopictus* vectors ^11^. In Brazil, dengue has been endemic for more than three decades in most of the country, except for the southern region where historically there was limited community transmission ^17,19^. In this study, we employed a combination of entomological and epidemiological analyses, including the virus transmission suitability index, alongside high-resolution genomic epidemiology, to investigate the historical spread of dengue in southern Brazil, particularly in Rio Grande do Sul, the southernmost Brazilian state. Our results demonstrated that dengue epidemiology has changed, driven by the complete *Ae. agypti* infestation across the state, followed by an increasing infestation level through the years. Moreover, overwintering endemic lineages are fuelling a yearly upsurge of cases in a region with an immunologically naive population. These findings have important implications for dengue mitigation in the region to reduce the overall burden, and on a global scale, as the increase of dengue in previously non-endemic areas is likely to occur in other temperate regions worldwide.

The marked spread of *Ae. aegypti* in the state, invading nearly all municipalities by 2023, is in line with previous forecast studies of higher suitability of subtropical regions for the vector due to climate change ^53^. Dengue incidence gradually increased in the East, North, and Northwest regions of the state, which coincide with the most densely populated regions. Harish et al. 2024, analyzing historical records of dengue in Brazil, estimated that a municipality is invaded by dengue when the incidence reaches 20 cases per 100,000 inhabitants. Based on this threshold, we detected a steady increase of invaded municipalities in Rio Grande do Sul, where almost half of the municipalities crossed the incidence threshold by 2023. However, we found no clear association of enlarged transmission seasons (more months with index P >1) in the state that could support a potential impact of the ongoing climate change in the region. It is important to note that climate change may be not captured within the nine-year time frame of the study and that the differential invasion of the vector and increase in case incidence between the temperate and tropical Brazil may be also explained by a delayed spread or adaptation of the vector to colder regions. Such open and important questions warrant further studies in the near future.

Our findings demonstrate a strong positive exponential correlation between the mosquito-borne viral suitability index (P index) and dengue incidence when a two-month lag is considered. Interestingly, the slope of this correlation (K constant) increased over the study period, particularly after 2021, suggesting that additional factors may have influenced dengue transmission dynamics. Given that this period coincided with the progressive geographic spread of DENV into previously naïve regions, we hypothesized that viral expansion played a role in shaping incidence patterns. Supporting this, linear regression analysis revealed a direct relationship between the K constant and the annual fraction of dengue-positive municipalities, indicating that the extent of viral spread within a given subregion is a key factor modulating incidence beyond vector suitability alone. These findings highlight the need for integrative models that account for both ecological and epidemiological factors in dengue forecasting.

The strong predictive performance of the two-month lagged P index underscores its potential utility for dengue forecasting. The ability to anticipate epidemic months with high sensitivity and specificity could support early warning systems and guide public health interventions. However, as dengue epidemiology evolves, the impact of acquired immunity—both from natural infections and vaccination—must be considered in predictive models. The progressive introduction of the dengue vaccine in Brazil, along with the increasing proportion of immune individuals in previously naïve regions, may alter transmission dynamics and reduce the predictive strength of historical correlations. Future studies should incorporate seroprevalence data and vaccine coverage rates to refine forecasting models, ensuring their continued reliability in the face of changing immunity landscapes.

Next, we performed phylogeographic analyses to evaluate if the increase in dengue cases in Rio Grande do Sul was linked to the continued introduction of new lineages or due to the endemic establishment and multi-year transmission chains. Our high-resolution genomic analysis showed multiple dengue lineages overwintering in Rio Grande do Sul. Additionally, the 2020–2023 outbreaks were majorly fueled by genotypes and lineages that were already circulating in the state in the previous years, supporting the endemic establishment of dengue lineages within the state. Lineage overwintering may occur either through vertical transmission of the virus to mosquito eggs or through asymptomatic cases, both of which can allow cryptic virus transmission in the region and a rapid increase in transmission during the next warm season, coinciding with the mosquito population surge ^54^. Endemic establishment and overwintering virus transmission means that no reintroduction of the virus is required for new outbreaks to occur.

The phylogeographic dynamics of DENV lineages within the state also were assessed. Most interstate transmission events during the non-endemic phase (2015–2020) occurred through the most populous Porto Alegre metropolitan region, with further intra-state virus spread to the Northeast, North, and Northwest regions, or through the Northwest region of the state to the Porto Alegre Metropolitan region. During the endemization phase (2021–2023), transmission chains were again detected in these more populous regions, in addition to multiple transmission chains at the Center-West, Center-East, and Southwest regions, covering the whole state, with DENV-1 1E-BR5 as the most prevalent lineage. These findings are consistent with the pattern of *Aedes* invasion in the state, with the first invaded municipalities concentrated in the Metropolitan and Northwest regions, compared to the more recently invaded municipalities in the South, Center-West, Center-East, and West regions of the state. Similarly, based on the year of invasion of the municipalities, those located in the Northeast, East, and Center-West regions were among the first to be invaded, whereas municipalities in the Northeast, Metropolitan, and Southwest regions were invaded later (Figure S15). Interestingly, municipalities from the Northwest and Metropolitan regions were the main transmission hubs within the state (Figures 6 and 7). The Northwest region, one of the earliest to be invaded, became an important source and sink hub, while the Metropolitan region, despite being invaded later, also emerged as a significant source and sink hub.

The DENV-1 1V_E lineage, responsible for almost 50% of the sequenced samples, shifted from multiple introductions between 2015 and 2019 to a more endemic behavior after 2020. This shift likely contributed to its prominent role in the smaller 2016 and larger 2022 outbreaks, spreading mainly from the Metropolitan region toward the central and northwestern areas. In contrast, the emerging DENV-1 1V_D.1 lineage, with a tMRCA in November 2020 and first detected in Peru in 2021, showed a rapid establishment in 2022, comprising more than 30% of the samples. Its unique diffusion pattern, moving from the Northwest region toward the Metropolitan region, highlights the heterogeneity in the spatial dynamics of DENV-1 lineages in Southern Brazil. The DENV-2 2II_F.1.2 lineage (Cosmopolitan genotype), which emerged more recently, followed a similar pattern of spread from the Metropolitan to the Northwest region, as seen in other DENV-1 lineages such as 1V_A, 1V_F, and 1V_E (BR5 and BR6), and 2III_C.1.1. The only exception to this pattern was DENV-1 1V_D.1, which spread in the opposite direction, from the Northwest to the Metropolitan region. These results emphasize the interplay between endemic persistence and recent introductions in shaping DENV transmission in the region, with important implications for the 2022 outbreak in particular.

The overall increasing dengue burden in Southern Brazil has far-reaching health consequences for the population. Accordingly, due to the historical absence or low prevalence of the virus in the region, the majority of the population is immunologically naive and, therefore, susceptible to dengue infection regardless of the circulating virus serotype, genotype, or lineage. This points to an alarming future with a rising incidence of dengue in the state. Additionally, the population in the southern region has a higher average age range compared to that of other Brazilian states ^55^. Although infection by one serotype of DENV generates immunity against another infection by the same serotype, a second infection by a different serotype is directly associated with a higher risk of DHF and DSS. Considering that different DENV serotypes circulate in Rio Grande do Sul, there is a greater risk of severe cases and even co-infection ^56^, especially in older individuals that are more impacted by comorbidities. Lastly, a recent study characterized that DENV serotype 3 is more associated with a higher incidence of symptomatic and severe cases in primary infections ^57,58^. DENV serotype 3 has recently reemerged in Brazil ^59^ and already spread to at least nine states ^60^, including Santa Catarina which borders Rio Grande do Sul to the north. Hence, we may anticipate more severe dengue cases in the future as DENV-3 continues to spread further south, particularly affecting the elderly and more vulnerable populations experiencing primary infections.

Our study has some limitations. First, genetic and genomic surveillance has been more systematically implemented in Brazil since 2023, but there are clear disparities between the states, hence the proportion of inter- and intra-state transmission chains must be evaluated with caution. This uneven sampling is evident in Supplementary Figure 16, which shows that, apart from 2019, sequencing efforts were largely concentrated in Rio Grande do Sul between 2020 and 2023. On the other hand, dengue molecular surveillance within Rio Grande do Sul state was implemented in a similar time frame as other Brazilian states, supporting that cases incidence in the state were mapped since the first cases and have increased throughout the years. Moreover, dengue genomic surveillance in the state is the most well-structured within Brazil where all samples that were above the threshold quality level were sequenced, meaning that we captured a substantial fraction of the DENV transmission chains. Second, as samples enter the surveillance pipeline through the healthcare facilities of the Brazilian Unified Health System, basically only symptomatic patients that seek health facilities are sampled. Therefore, our findings do not account for the asymptomatic dengue cases and their role in further transmission chains. Lastly, the large majority of the samples are registered in the system with very limited metadata regarding symptoms and disease manifestation, hence we were not able to assess the impact of disease severity and outcome of the different dengue lineages throughout the study period.

## Conclusion

Here, we characterize the increased dengue burden in subtropical Brazil through entomological and epidemiological analyses combined with high-resolution genomic information. This integrative analysis allowed us to associate the increasing dengue incidence to a combination of factore: the invasion and infestation level of the vector, geographical spread of dengue lineages and a susceptible population. From this, two key actionable insights emerged for mitigate dengue in the coming years: the identification of source and sink hubs within the state where control measures must be prioritized, and the use of the index P to forecast the size of an outbreak two months ahead of the peak in human cases. This early warning can help health care facilities scale up resources to support the affected population.

In Addition, these findings stress the urgent need to sustain dengue surveillance in the region, and also to improve and implement vector control measures along with more systematic implementation of vaccination campaigns to protect the overall population and the most vulnerable ones. Coordination between intersectoral investigations, mitigations, and clinical management approaches is imperative to limit the morbidity and mortality impact of dengue as it comes to infecting susceptible and vulnerable populations. Moreover, although our findings are restricted to a single subtropical region, the ongoing climate change and forecasting impact of vector-borne diseases strongly suggest that what we are witnessing in Southern Brazil will be reflected in other subtropical and temperate regions of the globe. Therefore, these findings and future dengue management in the region, may serve as an early indicator and a model for understanding dengue expansion and mitigation worldwide, especially as climate change continues to influence the spread of vector-borne diseases.

## Data Availability

All data produced in the present work are contained in the manuscript - Figshare.

## Data Availability

All the data needed to reproduce the findings of this manuscript are available through GISAID and Figshare (the link will be available as soon as it goes to the review stage).

## Ethics Approval

This project was approved by the Research Ethics Committee (CEP) at Escola de Saúde Pública (SES-RS). Process number: CAAE: 67181123.1.0000.5312

## Acknowledgement

We would like to thank all the public health professionals and researchers that are constantly battling against Dengue including health care professionals and researchers committed to sharing information to tackle this still neglected infectious disease.

