## Supplementary Files for "Increasing dengue outbreaks in temperate Brazil is linked to *Aedes aegypti* invasion and infestation level driving widespread virus transmission"

**This PDF file includes:**

Methods
Figs. S1 to S16
Tables S1 to S7
References

**Other Supplementary Material for this manuscript includes the following:**

Figshare:.....

Supplementary_Tables.xlsx (supplementary table 1, 4 and 7)

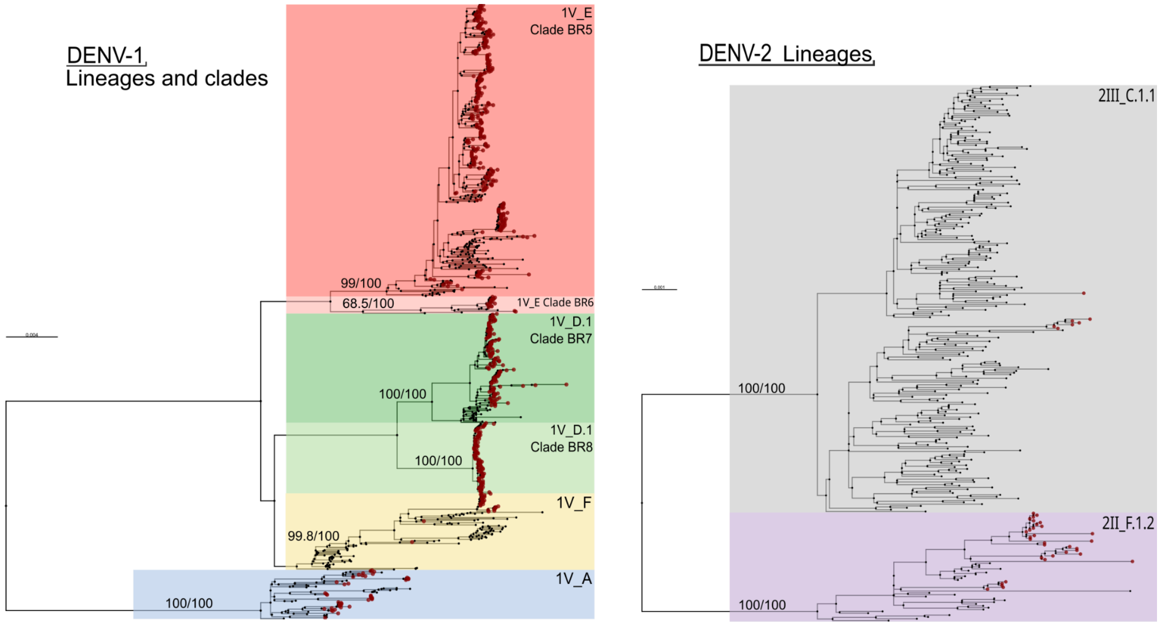


Figure S1. Maximum likelihood (ML) phylogenetic trees of DENV-1 (left) and DENV-2 (right), based on a global dataset. The original trees included 3,476 DENV-1 and 3,091 DENV-2 genomes, but here they are shown in a collapsed format, retaining only the clades and lineages that contain our study samples. In this collapsed representation, the DENV-1 tree comprises 773 genomes, while the DENV-2 tree includes 280 genomes. In the DENV-1 tree, the six identified clades are highlighted with different colors and labeled in the upper-right corner of each highlighted region. Similarly, in the DENV-2 tree, the identified lineages are highlighted and labeled accordingly. Bootstrap support values are shown at key nodes leading to major clades. Tip points in red represent our study samples, while black tip points correspond to sequences retrieved from NCBI.

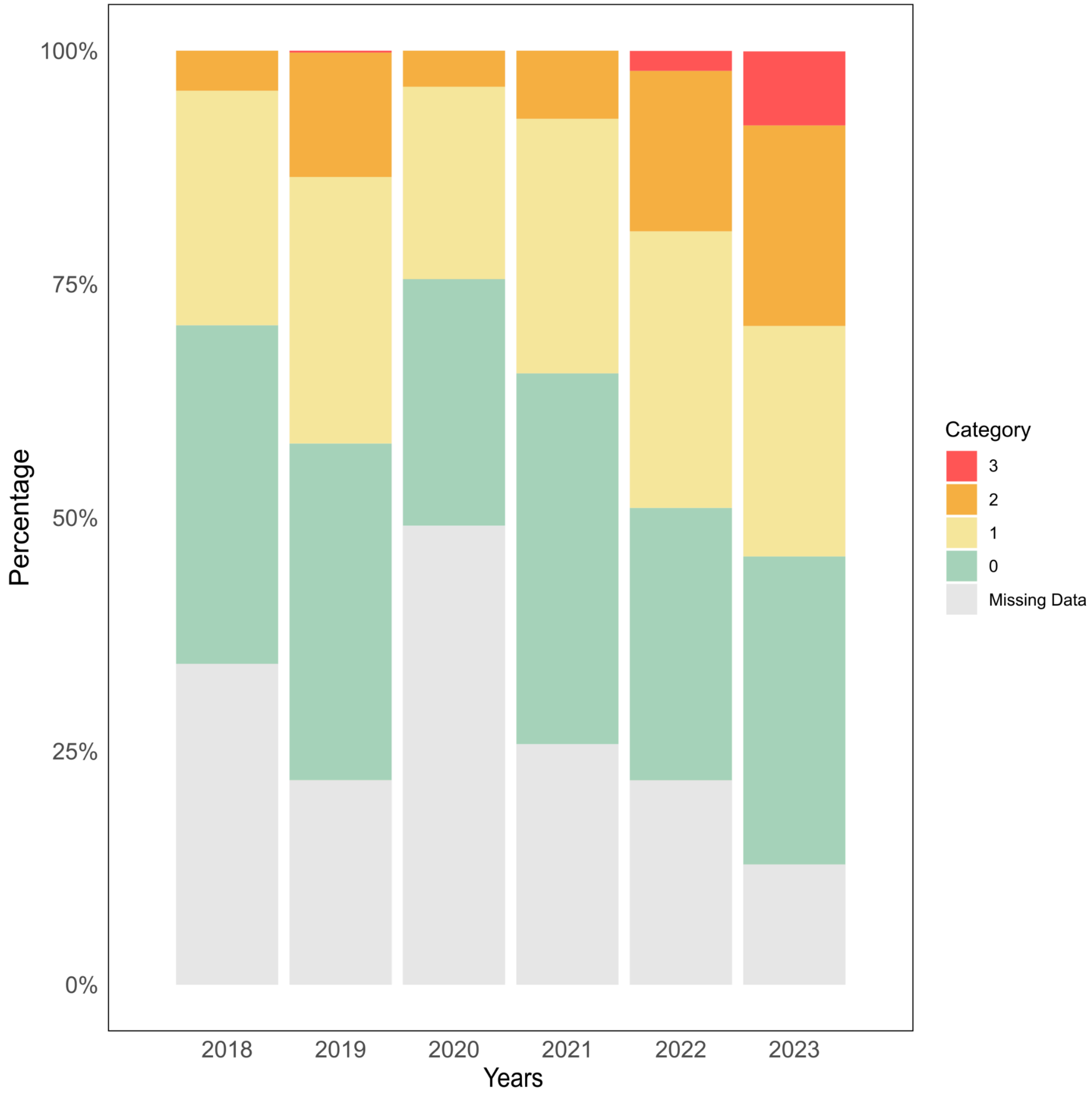


Figure S2 - Annual distribution of *Aedes aegypti* infestation levels in Rio Grande do Sul from 2018 to 2023. The classification follows the Brazilian Ministry of Health criteria: 0 (absence of infestation, green), 1 (low infestation, yellow), 2 (moderate infestation, orange), and 3 (high infestation, red). Missing data are represented in gray. The increasing proportion of municipalities in categories 2 and 3 over time highlights the expansion of infestation in the state.

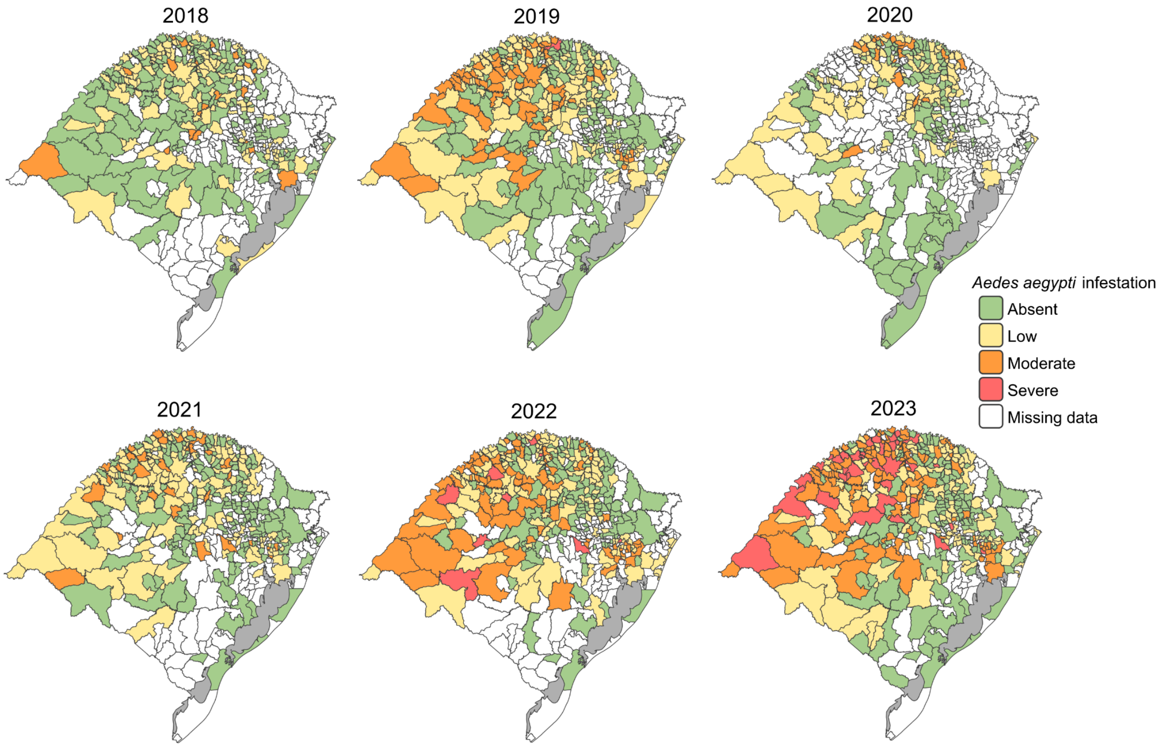


Figure S3. Spatial distribution of *Aedes aegypti* infestation levels in Rio Grande do Sul from 2018 to 2023. Each map represents the classification of municipalities based on the Brazilian Ministry of Health criteria: 0 (absence of infestation, green), 1 (low infestation, yellow), 2 (moderate infestation, orange), and 3 (high infestation, red). Municipalities with missing data are shown in white. The maps illustrate the expansion of infestation over time, with a growing number of municipalities classified in the moderate and high infestation categories.

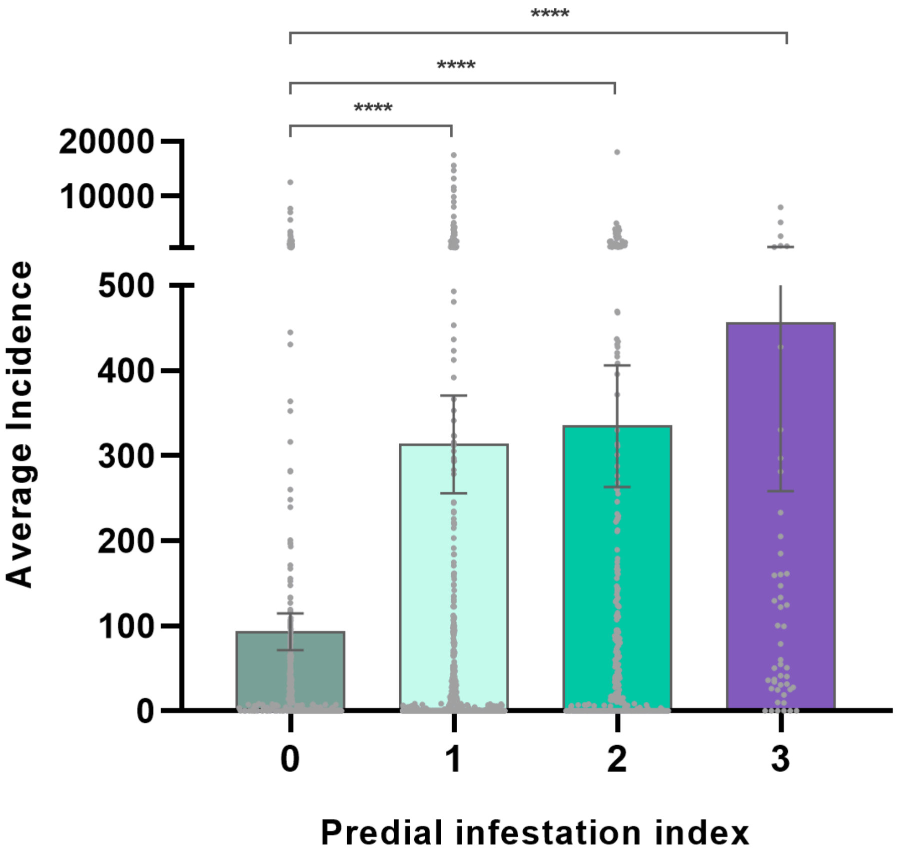


Figure S4. Association between the *Aedes aegypti* infestation index and average dengue incidence (2018–2023) in municipalities of Rio Grande do Sul. Municipalities were classified into four infestation levels (LIRA 0–3). Bars represent mean dengue incidence (cases per 100,000 inhabitants), with individual municipality values shown as dots. Significant differences between groups are indicated ($p < 0.0001$ for LIRA 1, 2, and 3 vs. 0; $p = 0.0014$ for LIRA 3 vs. 2)

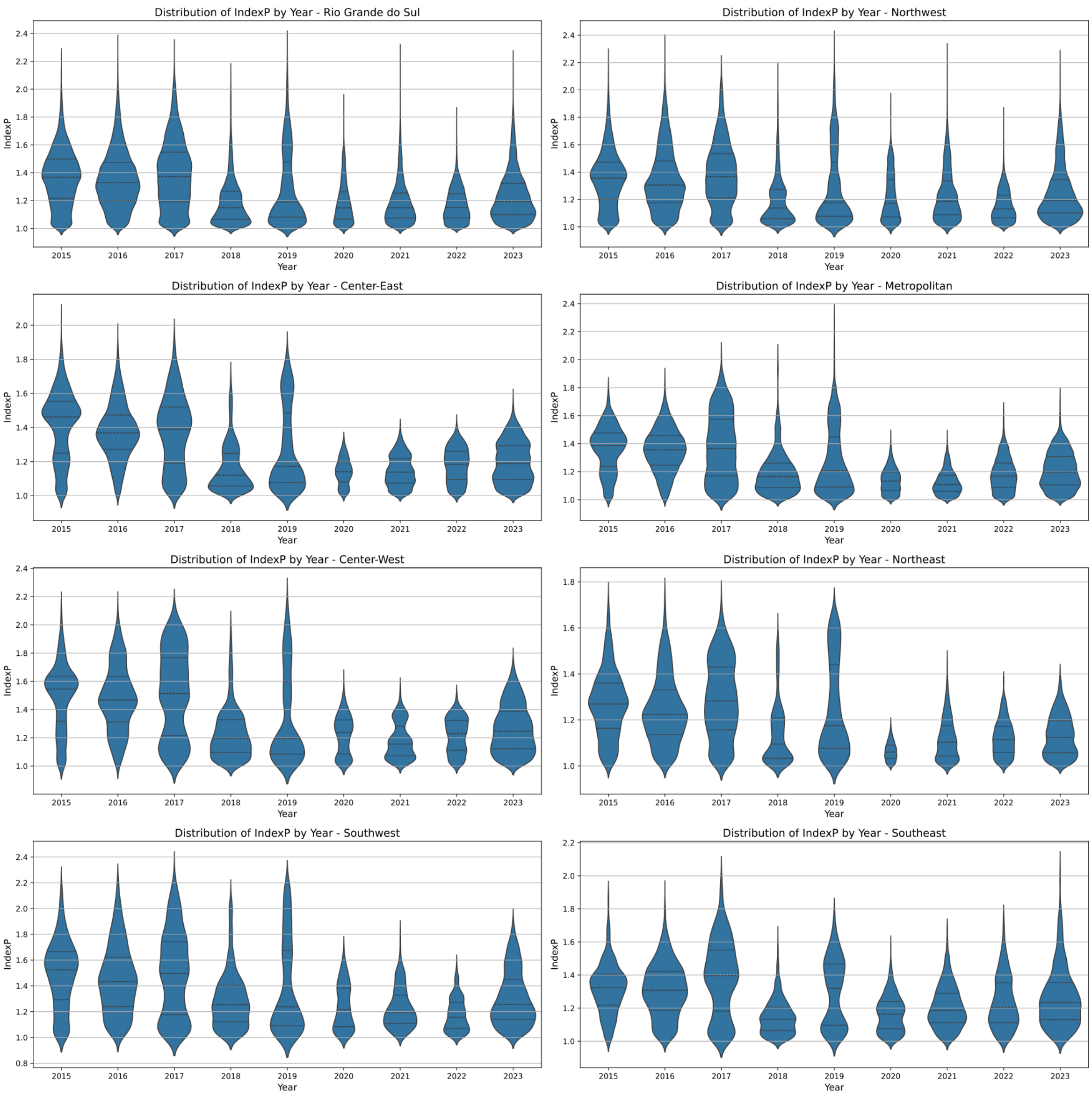


Figure S5. Temporal variation of the mosquito-borne viral suitability index (*Index P*) across mesoregions of Rio Grande do Sul from 2015 to 2023. Each panel represents a different mesoregion, with an additional panel for the entire state. The x-axis corresponds to the months of the year, while the left y-axis represents the years analyzed. The right y-axis shows the *Index P* distribution, represented as violin plots. These results highlight the seasonal pattern of suitability, with higher values typically observed between January and March.

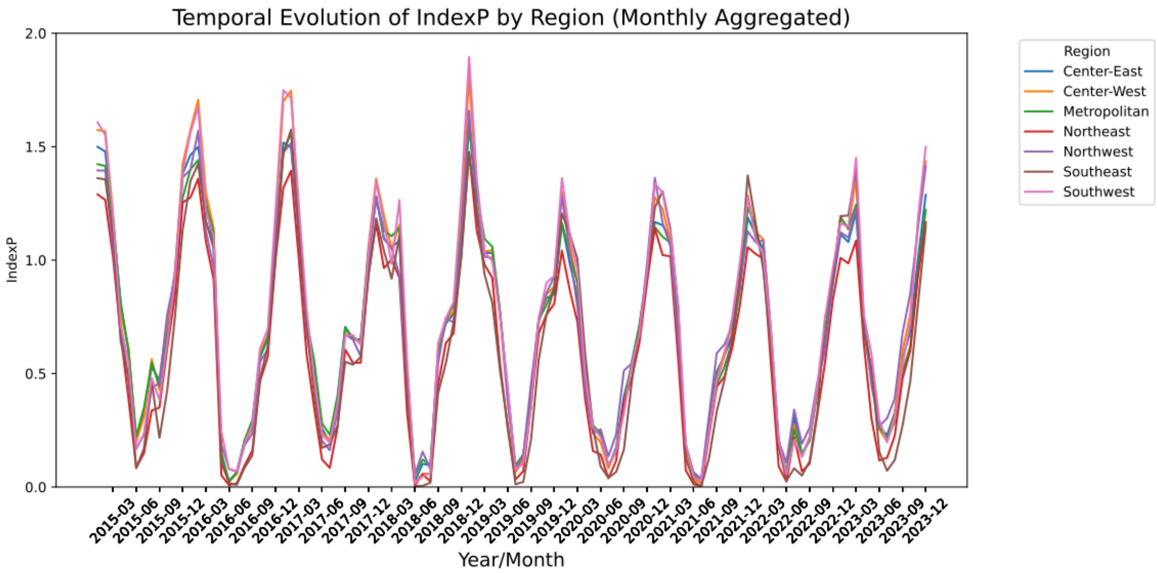


Figure S6. Temporal variation of the mosquito-borne viral suitability index (Index P) across mesoregions of Rio Grande do Sul from 2015 to 2023. This figure presents the aggregated results for all mesoregions, with each region represented by a distinct line color. The x-axis corresponds to the months and years analyzed, while the y-axis shows the Index P values. The overlapping trends illustrate the overall seasonal pattern, with higher suitability typically observed between January and March.

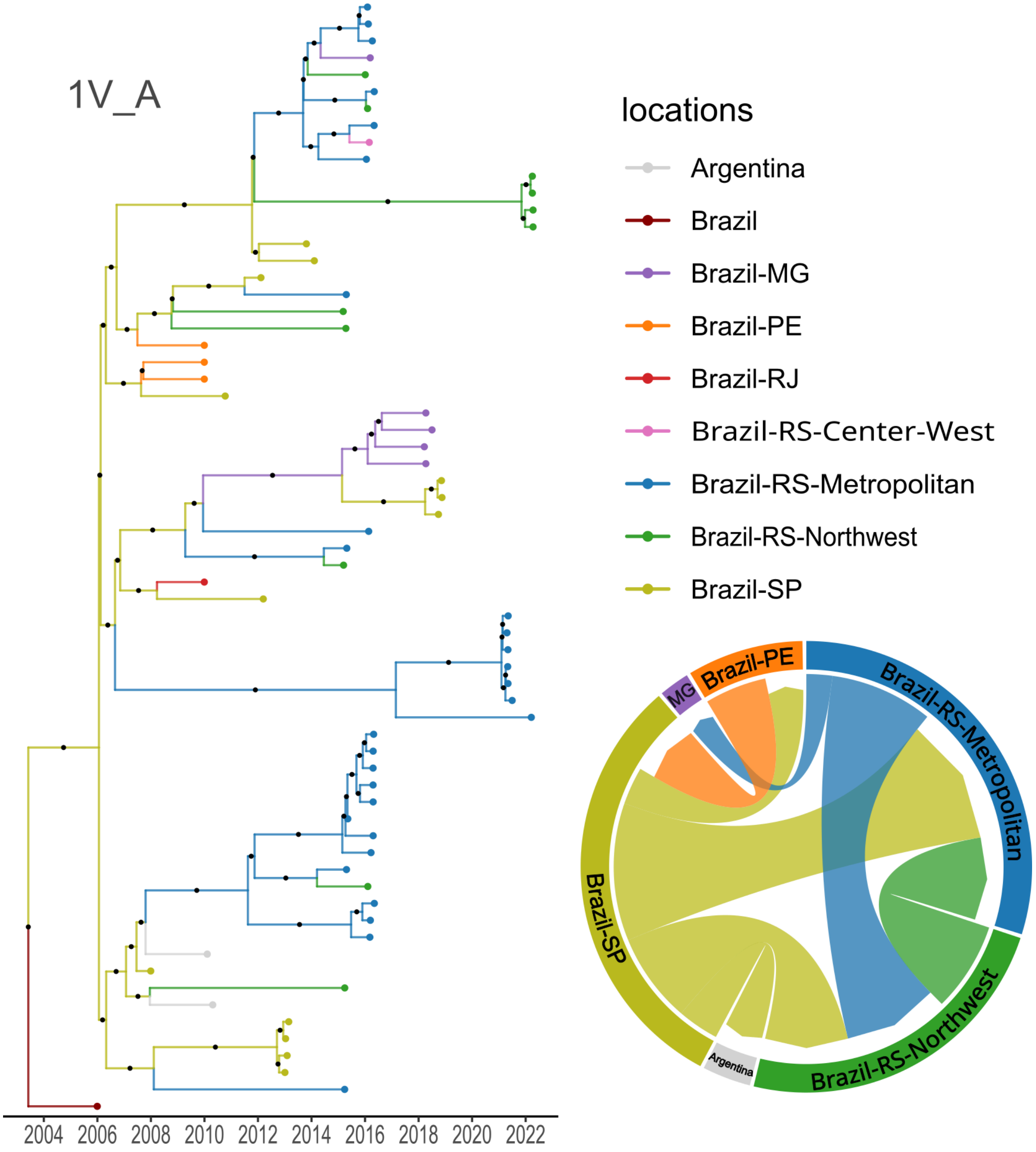


Figure S7. Time-scaled maximum clade credibility (MCC) tree and circular migration plot of the DENV-1 lineage 1V_A. The MCC tree represents the evolutionary history of the lineage, with branch lengths proportional to time and color gradients indicating inferred geographic transitions. The circular migration plot, derived from Markov jump analyses, illustrates the frequency and directionality of viral dispersal between different regions of Rio Grande do Sul.

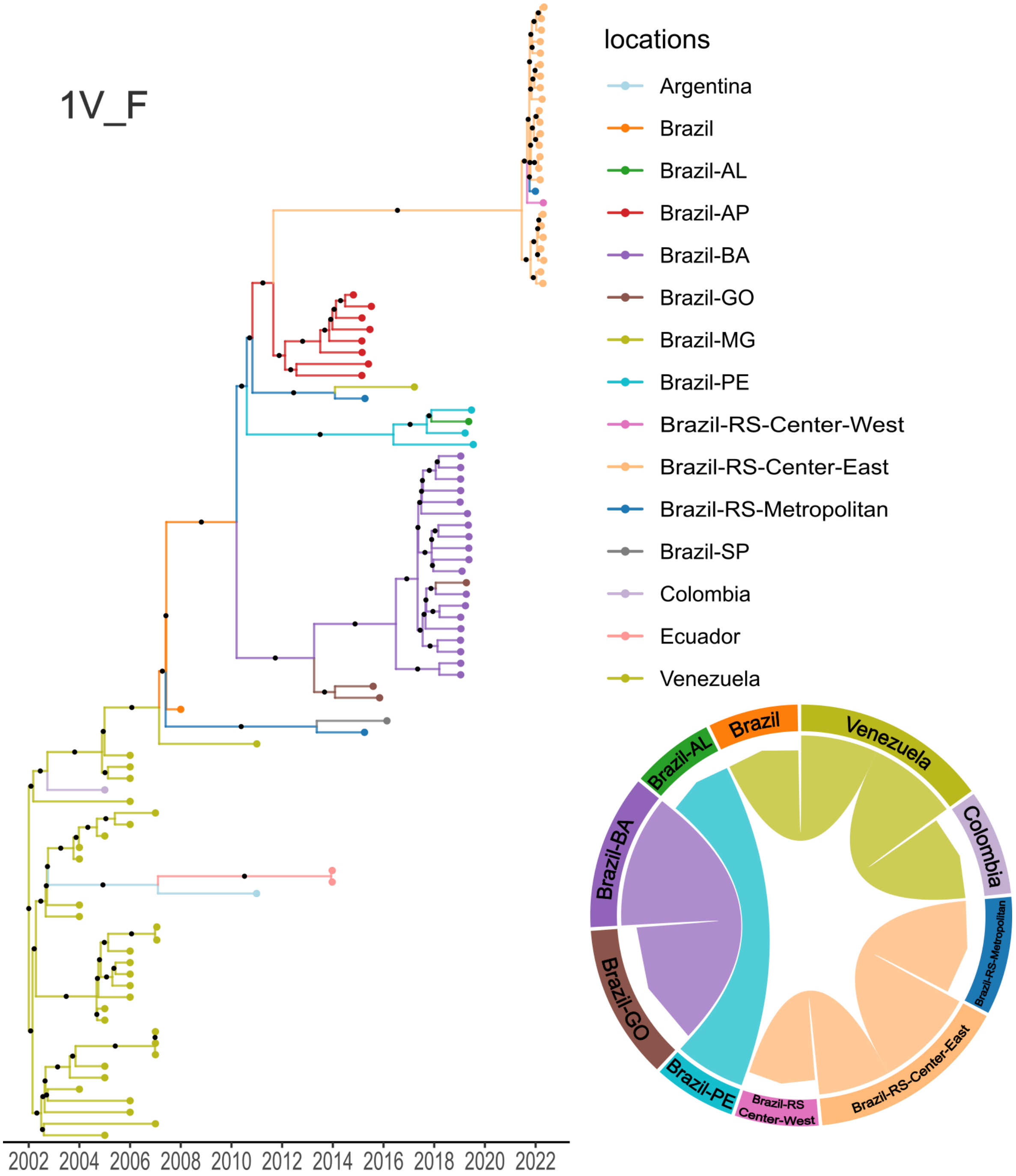


Figure S8. Time-scaled maximum clade credibility (MCC) tree and circular migration plot of the DENV-1 lineage 1V_F. The MCC tree represents the evolutionary history of the lineage, with branch lengths proportional to time and color gradients indicating inferred geographic transitions. The circular migration plot, derived from Markov jump analyses, illustrates the frequency and directionality of viral dispersal between different regions of Rio Grande do Sul.

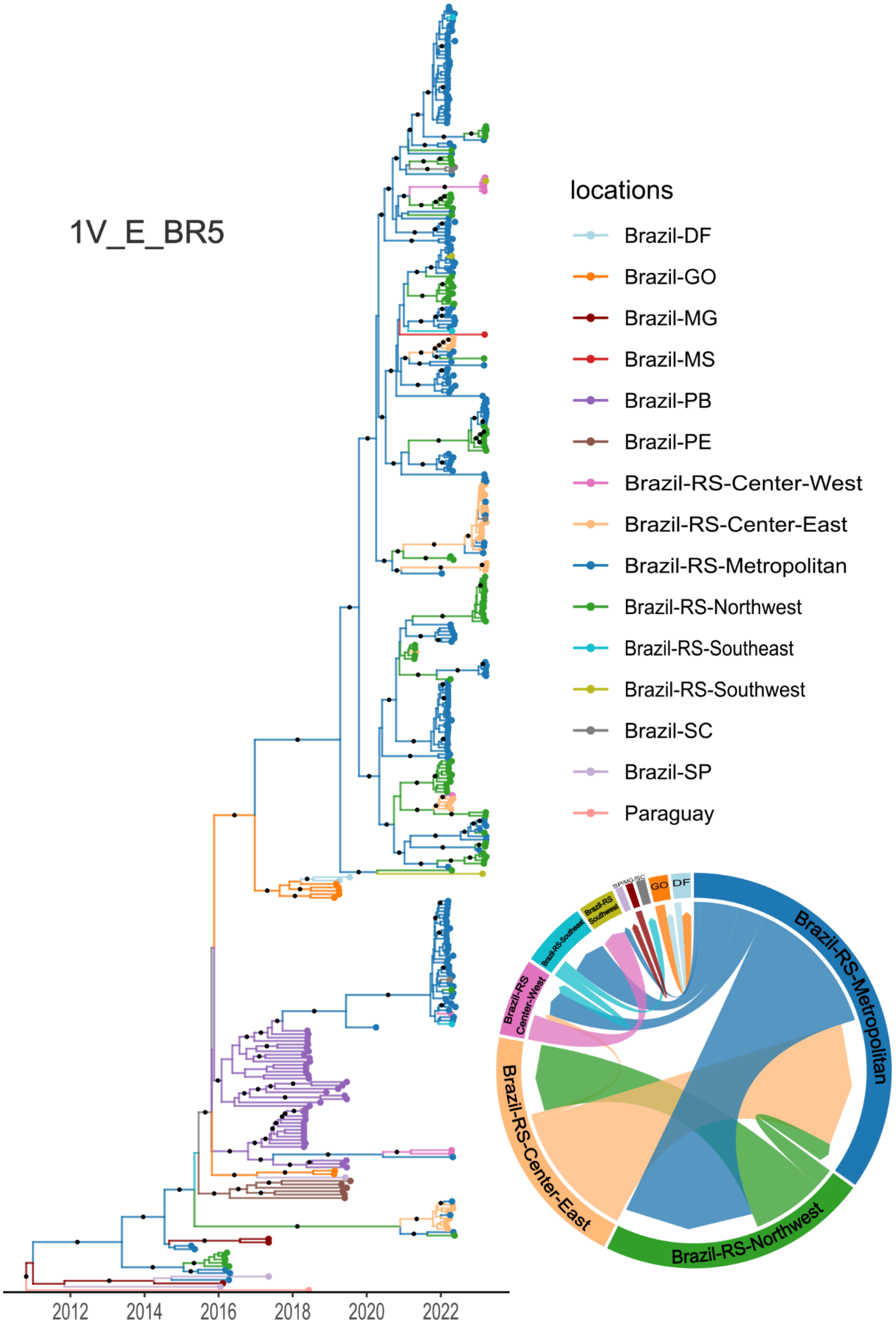


Figure S9. Time-scaled maximum clade credibility (MCC) tree and circular migration plot of the DENV-1 lineage 1V_E_BR5. The MCC tree represents the evolutionary history of the lineage, with branch lengths proportional to time and color gradients indicating inferred geographic transitions. The circular migration plot, derived from Markov jump analyses, illustrates the frequency and directionality of viral dispersal between different regions of Rio Grande do Sul.

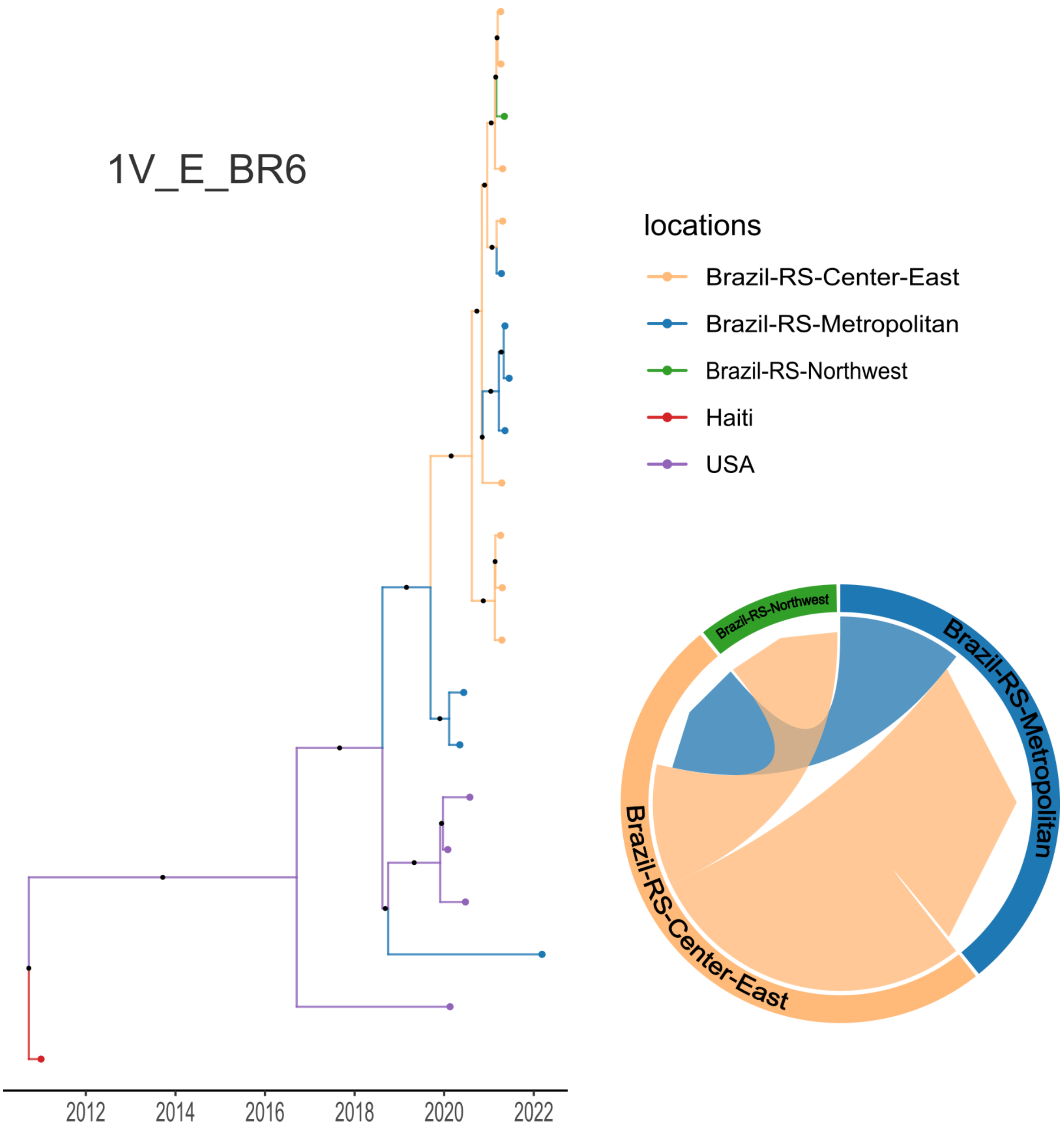


Figure S10. Time-scaled maximum clade credibility (MCC) tree and circular migration plot of the DENV-1 lineage 1V_E_BR6. The MCC tree represents the evolutionary history of the lineage, with branch lengths proportional to time and color gradients indicating inferred geographic transitions. The circular migration plot, derived from Markov jump analyses, illustrates the frequency and directionality of viral dispersal between different regions of Rio Grande do Sul.

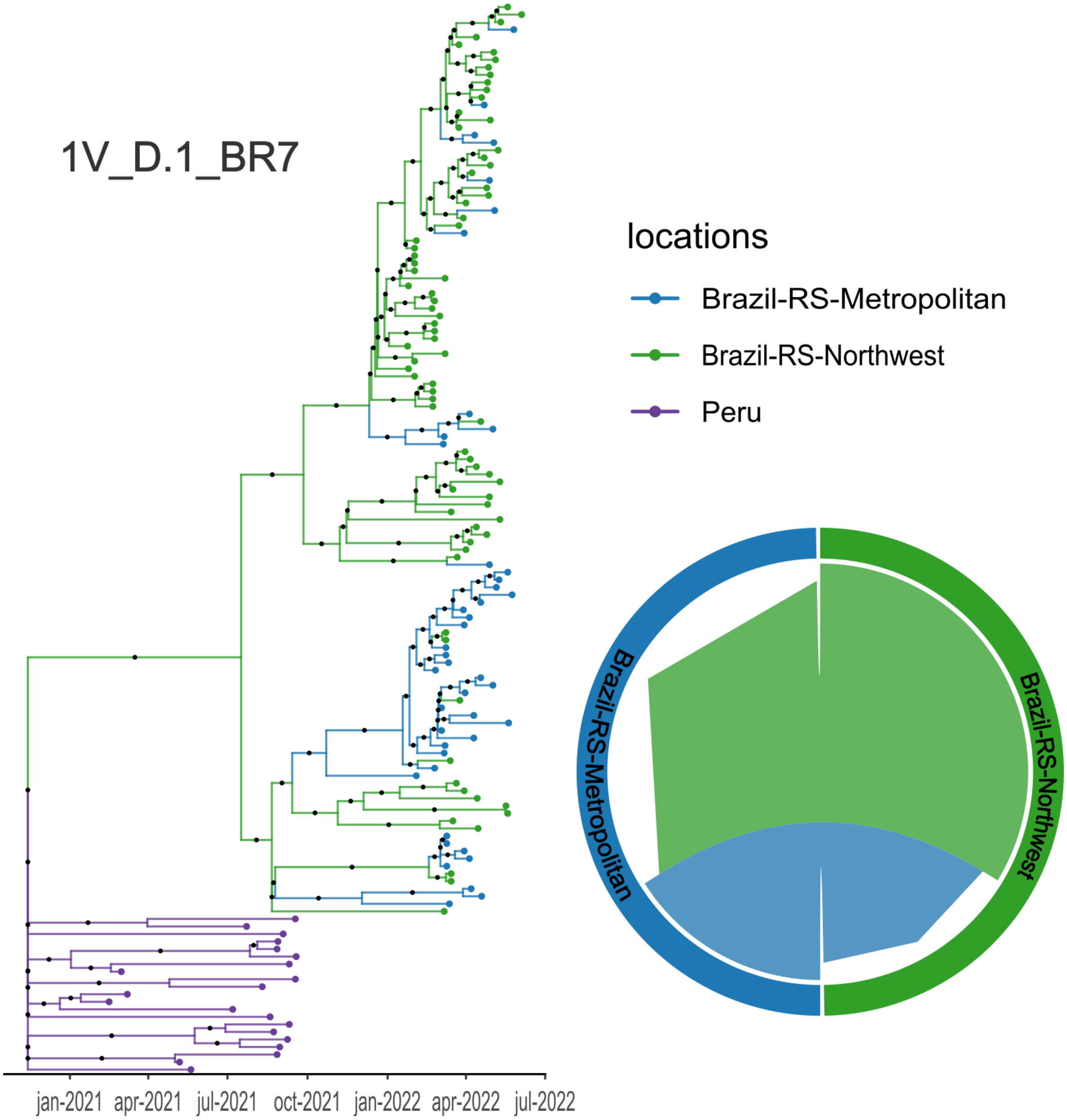


Figure S11. Time-scaled maximum clade credibility (MCC) tree and circular migration plot of the DENV-1 lineage 1V_D.1_BR7. The MCC tree represents the evolutionary history of the lineage, with branch lengths proportional to time and color gradients indicating inferred geographic transitions. The circular migration plot, derived from Markov jump analyses, illustrates the frequency and directionality of viral dispersal between different regions of Rio Grande do Sul.

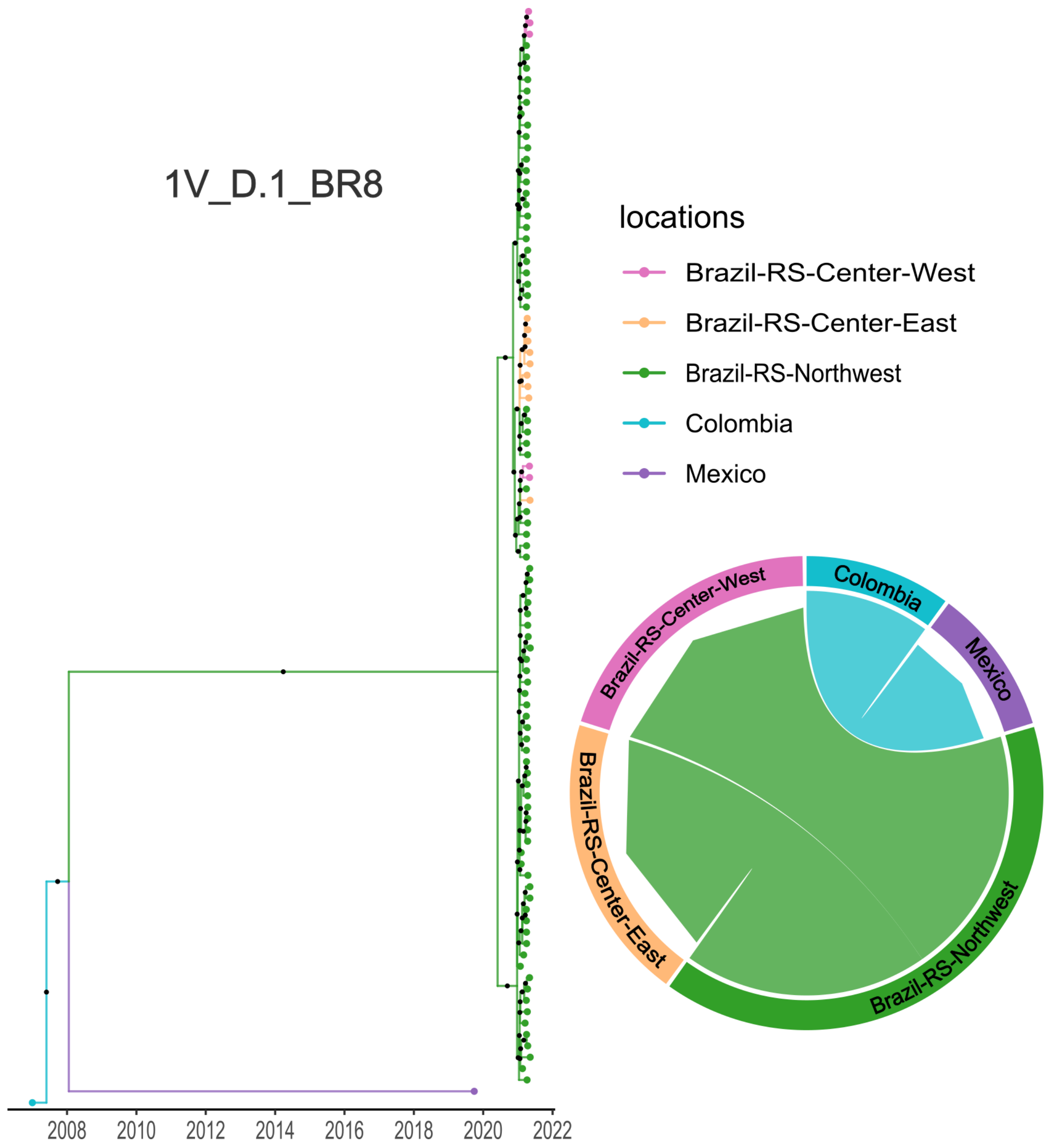


Figure S12. Time-scaled maximum clade credibility (MCC) tree and circular migration plot of the DENV-1 lineage 1V_D.1_BR8. The MCC tree represents the evolutionary history of the lineage, with branch lengths proportional to time and color gradients indicating inferred geographic transitions. The circular migration plot, derived from Markov jump analyses, illustrates the frequency and directionality of viral dispersal between different regions of Rio Grande do Sul.

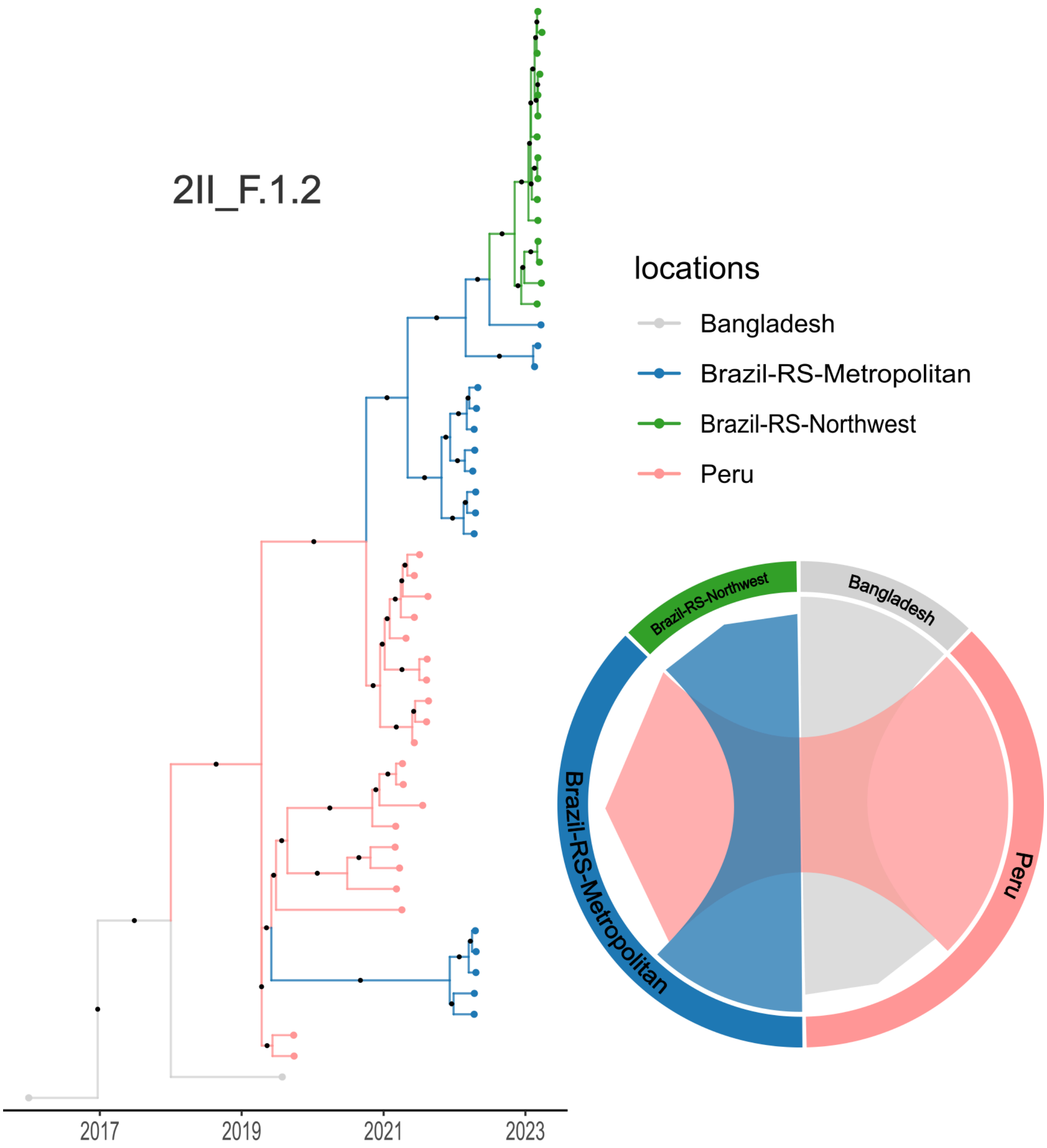


Figure S13. Time-scaled maximum clade credibility (MCC) tree and circular migration plot of the DENV-2 lineage 2II_F.1.2. The MCC tree represents the evolutionary history of the lineage, with branch lengths proportional to time and color gradients indicating inferred geographic transitions. The circular migration plot, derived from Markov jump analyses, illustrates the frequency and directionality of viral dispersal between different regions of Rio Grande do Sul.

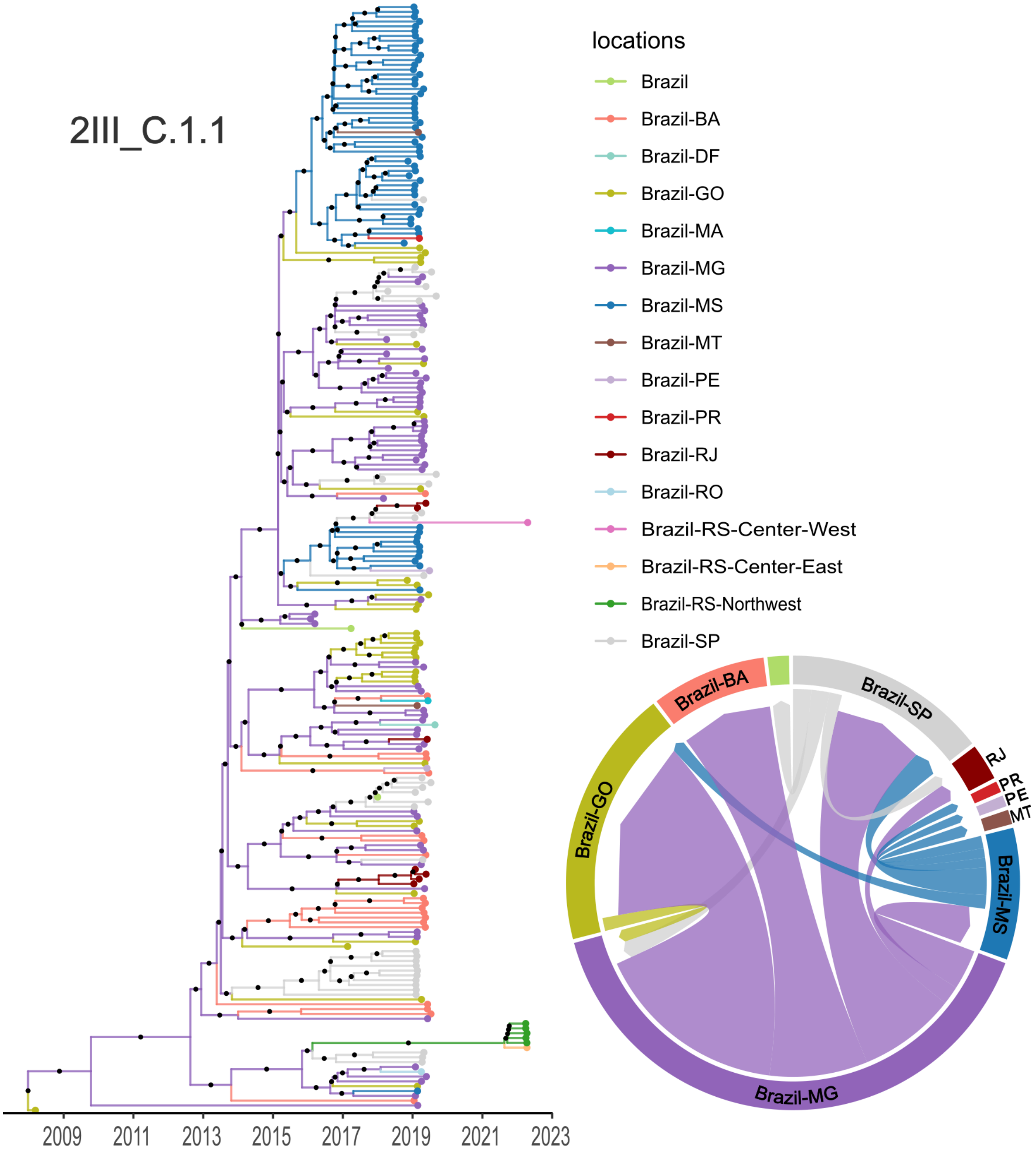


Figure S14. Time-scaled maximum clade credibility (MCC) tree and circular migration plot of the DENV-2 lineage 2III_C.1.1. The MCC tree represents the evolutionary history of the lineage, with branch lengths proportional to time and color gradients indicating inferred geographic transitions. The circular migration plot, derived from Markov jump analyses, illustrates the frequency and directionality of viral dispersal between different regions of Rio Grande do Sul.

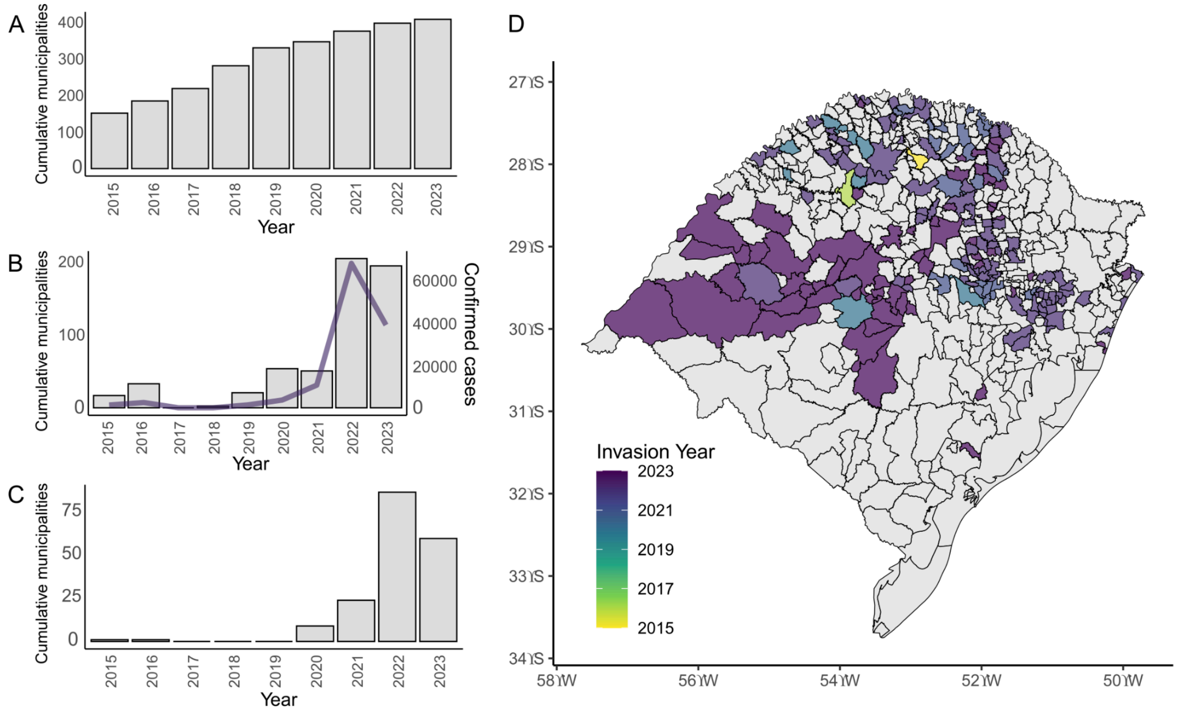


Figure S15. *Aedes aegypti* invasion and dengue virus spread in Rio Grande do Sul. (A) Number of municipalities considered infested by the *Aedes* mosquito per year. (B) Number of municipalities showing signs of dengue invasion (an incidence of 20 or more cases per 100,000 inhabitants per year). (C) Number of municipalities with dengue emergence and subsequent persistence per year. (D) Map of the state of Rio Grande do Sul showing the year of dengue emergence in each municipality.

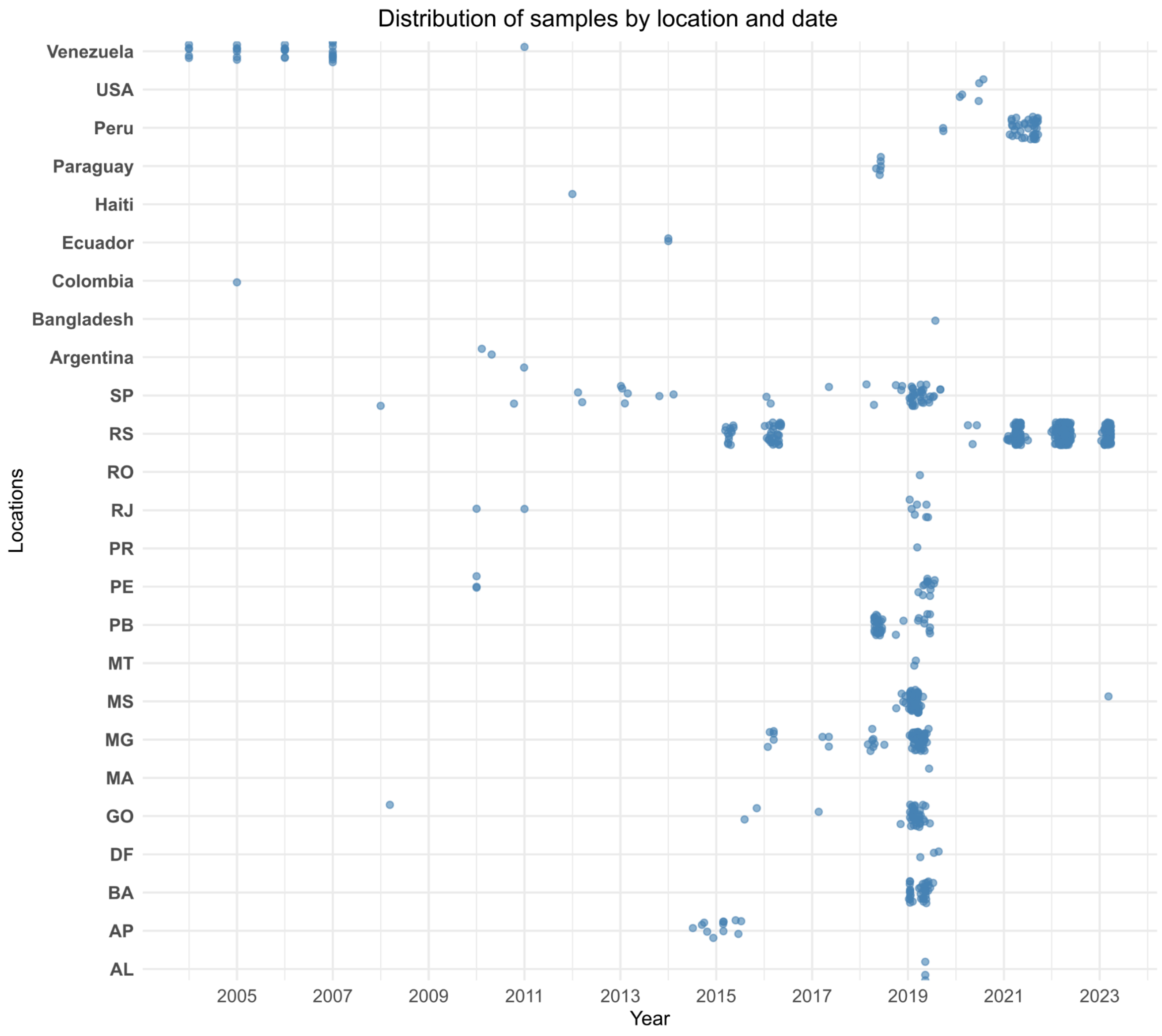


Figure S16. Distribution of genome sampling by location and year. The y-axis represents the sampling locations, while the x-axis shows the collection dates. The dataset shows an uneven distribution of sequences across locations and time, with certain regions and periods having greater representation, which influences the scope of the phylodynamic analysis.

**Supplementary Table 1**. Genome Assembly Quality Metrics for DENV-1 and DENV-2 Samples. This table summarizes the genome assembly quality metrics for DENV-1 samples analyzed in this study. The columns include the sample identifier (cod), mean sequencing depth (mean_depth_coverage), standard deviation of sequencing depth (sd_depth_coverage), median sequencing depth (median_depth_coverage), and genome coverage breadth (coverage_breadth).

Supplementary_tables.xlsx - sheet: Supplementary table 1

Supplementary table 2. DENV-1 and DENV-2 subsets used for phylodynamic analysis. The table presents the number of new sequenced genomes, the total genomes per subset, the number of removed outliers, the final number of genomes after outlier removal, the subset frequency representing the proportion of each subset relative to the total number of genomes, and the correlation coefficient indicating the strength of the temporal relationship observed between genetic distance and sample collection dates.

| **DENV-1 subsets** | **New Genomes** | **Total Genomes** | **Outliers Removed (TempEst)** | **Final Genomes** | **Subset Frequency** | **Correlation Coefficient** |
|---|---|---|---|---|---|---|
| 1V_A | 40 | 66 | 0 | 66 | 6,015% | 0.88 |
| 1V_F | 27 | 99 | 4 | 95 | 4,06% | 0.95 |
| 1V_E_BR5 | 321 | 377 | 16 | 361 | 48,27% | 0.51 |
| 1V_E_BR6 | 17 | 21 | 2 | 19 | 2,556% | 0.98 |
| 1V_D.1_BR7 | 126 | 142 | 6 | 136 | 18,947% | 0.93 |
| 1V_D.1_BR8 | 95 | 102 | 6 | 96 | 14,285% | 0.97 |
| **DENV-2 Subsets** | **New Genomes** | **Total Genomes** | **Outliers Removed (TempEst)** | **Final Genomes** | **Subset Frequency** | **Correlation Coefficient** |
| 2II_F.1.2 | 32 | 53 | 2 | 51 | 4,81% | 0.84 |
| 2III_C.1.1 | 7 | 230 | 1 | 229 | 1,05% | 0.52 |

Supplementary table 3. Skygrid parameters for each of the 8 subsets of DENV-1 and DENV-2 analyzed in BEAST. This table summarizes the Skygrid model parameters used in the phylodynamic analysis for each subset. The sample size represents the number of sequences included in the analysis, while the empirical age range denotes the span of years covered by these samples. The root-to-tip age corresponds to the time range from the estimated root of the phylogenetic tree to the most recent sample. The estimated root age was inferred from the BEAST analysis. The time at the last transition point indicates the most recent time at which a demographic change was inferred, based on a separate skyline analysis. The number of grids defines the temporal resolution of the Skygrid model, with grid intervals tailored to each subset to optimize model performance. The grid justification column specifies the rationale for the chosen grid density, which varies based on the sampling time span and the demographic dynamics of each lineage.

| **Subset** | **1V_E_ BR5** | **1V_E_ BR6** | **1V_D.1 _BR8** | **1V_F** | **1V_A** | **1V_D.1 _BR7** | **2II_F.1.2** | **2III_C.1.1** |
|---|---|---|---|---|---|---|---|---|
| **Sample size (empirical)** | 377 | 21 | 97 | 99 | 66 | 142 | 53 | 230 |
| **Age range (empirical)** | 8 | 10 | 2 | 18 | 16 | 13 | 6 | 6 |
| **Root-to-tip age (empirical)** | 2015/ 2023 | 2012/ 2022 | 2004/ 2022 | 2004/ 2022 | 2006/ 2022 | 2021/ 2022 | 2017/ 2023 | 2016/ 2022 |
| **Estimated root age** | 2011 | 2008 | 2004 | 2002 | 2004 | 2020 | 2017 | 2008 |
| **Time at last transition point** | 12 | 14 | 18 | 20 | 18 | 15 | 7 | 8 |
| **Grids** | 48 | 14 | 18 | 40 | 36 | 30 | 14 | 32 |
| **Grid justification** | 4 grids per year | 1 grid per year | 1 grid per year | 2 grids per year. | 2 grids per year. | 2 grids per year. | 2 grids per year. | 4 grids per year |

**Supplementary table 4.** Model Comparison Using Path Sampling and Stepping-Stone Methods. Log marginal likelihood values estimated using path sampling (PS) and stepping-stone (SS) methods for different clock and demographic models in BEAST. Comparisons were made for six test groups (G1–G6), evaluating strict and relaxed molecular clocks with constant size, skyline, and skygrid demographic models.

Supplementary_tables.xlsx - sheet: Supplementary table 4

**Supplementary table 5.** ROC Curve results for P index as a predictor of a dengue epidemic month (2015-2023)

| | **2-months lag (95% CI)** | **No lag** |
|---|---|---|
| **AUC** | 0.91 | 0.69 |
| **Sensitivity** | 93,55 (79,28 - 98,85) | 58,06 (40,77 - 73,58) |
| **Specificity** | 84,00 (74,08 - 90,60) | 67,53 (56,46 - 76,94) |
| **Best cut-off value** | > 0.7426 | |

**Supplementary table 6.** Links to interactive spatial diffusion visualizations for all eight analyzed subsets, generated using SpreaD4 (https://spreadviz.org/home). These visualizations provide an interactive representation of the inferred dispersal routes and geographic spread of each lineage in Rio Grande do Sul.

| Subset | Link |
|---|---|
| 1V_A | https://view.spreadviz.org/?output=c8e8b3a5-00ad-480a-980c-553b54e79570/458e206e-2cf7-4ba8-9aa2-3b085db6d4c4.json&maps=AR,PY,UY,BO,BR |
| 1V_F | https://view.spreadviz.org/?output=c8e8b3a5-00ad-480a-980c-553b54e79570/e0e5d128-f02d-4ad2-9fec-ee6cde7b2051.json&maps=CO,AR,VE,SR,PY,GY,UY |
| 1V_E_BR5 | https://view.spreadviz.org/?output=c8e8b3a5-00ad-480a-980c-553b54e79570/e3c828c0-99b6-400e-8eab-a035de4e134a.json&maps=AR,PY,UY,BO,BR |
| 1V_E_BR6 | https://view.spreadviz.org/?output=c8e8b3a5-00ad-480a-980c-553b54e79570/64acbe1e-1eda-4e14-9b07-8a06c0f8e8d4.json&maps=CO,PR,AG,VI,BQ,AW,LC |
| 1V_D.1_BR7 | https://view.spreadviz.org/?output=c8e8b3a5-00ad-480a-980c-553b54e79570/91566c13-6d36-4c14-8401-9a55bd49fceb.json&maps=AR,PY,UY,BO,CL,BR,PE |
| 1V_D.1_BR8 | https://view.spreadviz.org/?output=c8e8b3a5-00ad-480a-980c-553b54e79570/7a27ba43-d97f-4440-ac1c-0b03336f3eb2.json&maps=CO,PR,AG,VI,BQ,AW,LC |
| 2II_F.1.2 | https://view.spreadviz.org/?output=c8e8b3a5-00ad-480a-980c-553b54e79570/6641616f-3988-4527-8e0a-428809f4164e.json&maps=CF,SO,CO,PR,SS,AO,AG |
| 2III_C.1.1 | https://view.spreadviz.org/?output=c8e8b3a5-00ad-480a-980c-553b54e79570/923618c6-a899-4736-8777-aee61ba382c8.json&maps=AR,PY,BO,BR |

**Supplementary table 7.** Well-Supported Regional Transition Events Inferred from Markov Jumps. This table presents the well-supported transition events between geographic regions inferred from Markov jump analyses for each subset. Rows represent the origin of the transition, while columns indicate the destination. The values correspond to the number of well-supported transitions between regions, with higher values suggesting more frequent or strongly supported movement. Zero values indicate no significant transition detected between the respective regions.

Supplementary_tables.xlsx - sheet: Supplementary table 7
